# Time to negative throat culture following initiation of antibiotics for pharyngeal group A *Streptococcus*: a systematic review and meta-analysis to inform public health control measures

**DOI:** 10.1101/2022.11.08.22282068

**Authors:** Emma McGuire, Ang Li, Simon M Collin, Valerie Decraene, Michael Cook, Simon Padfield, Shiranee Sriskandan, Chris Van Beneden, Theresa Lamagni, Colin S Brown

## Abstract

**Background:** Public health guidance recommending isolation of individuals with group A streptococcal (GAS) infection or carriage for 12-24 hours from antibiotic initiation to prevent onward transmission requires a strong evidence-base.

**Methods:** We conducted a systematic review (PROSPERO CRD42021290364) and meta-analysis to estimate the pooled proportion of individuals who remain GAS culture-positive at set intervals after initiation of antibiotics. We searched Ovid MEDLINE (1946-), EMBASE (1974-) and the Cochrane library. We included interventional or observational studies with ten or more participants reporting rates of GAS throat culture during antibiotics for culture confirmed GAS pharyngitis, scarlet fever, and asymptomatic pharyngeal GAS carriage. We did not apply age, language, or geographical restrictions.

**Findings:** Of 5,058 unique records identified, 43 were included; 37 (86%) randomised controlled studies, three (7%) non-randomised controlled trials and three (7%) before-and-after studies. The proportion of individuals who remained culture-positive at day 1, day 2, and day 3-9 were 6.9% (95% CI 2.7-16.8%), 5.4% (95% CI 2.1-13.3%) and 2.6% (95% CI 1.6-4.2%). For penicillins and cephalosporins, day 1 positivity was 6.5% (95% CI 2.5-16.1%) and 1.6% (95% CI 0.04-42.9%) respectively. Overall, for 9.1% (95% CI 7.3-11.3), throat swabs collected after completion of therapy were GAS culture-positive.

**Interpretation:** Our review provides evidence that antibiotics for pharyngeal GAS achieve a high rate of culture conversion within 24 hours but highlights the need for further research given the methodological limitations of published studies and imprecision of pooled estimates. Further evidence is needed for non-beta-lactam antibiotics and for asymptomatic individuals.

## INTRODUCTION

*Streptococcus pyogenes* (group *A Streptococcus*, GAS) causes a range of clinical syndromes from mild infections such as impetigo, pharyngitis and scarlet fever to life-threatening invasive group A streptococcal (iGAS) infection. Severe infections predominantly affect the elderly, infants, and women who have recently given birth [1]. The prevalence of asymptomatic GAS throat carriage is approximately 7% (95% CI 5.6-8.8%) globally with higher rates reported in children (8%, 95% CI 6.6-9.7%) [2]. However, reported rates rarely exceed 1% in the UK [3, 4].

Outbreaks arise from both symptomatic and asymptomatic index cases and can occur in a range of settings including households, hospitals, and care homes [5-7]. Without treatment, the secondary attack rate for pharyngitis and scarlet fever is estimated to be 25-35% within a family setting and 23-60% in educational settings [8-10]. Even with treatment of the index case, attack rates among contacts are high (26% among school contacts, 13% among household contacts) [5]. In classroom outbreaks, prevalence of the outbreak strain has been noted to increase rapidly among asymptomatic contacts, reaching 27% in week two [5]. Within households, the median time between primary and secondary cases is six days [11]. GAS infections are treated with antibiotics to shorten duration of symptoms and reduce the risk of severe disease or long-term sequelae, including rheumatic fever [12, 13]. In GAS outbreaks, antibiotic treatment is recommended for individuals both with disease and with asymptomatic carriage to induce bacteriological clearance and prevent onward transmission.

Current public health guidance recommends that patients with GAS pharyngitis or scarlet fever stay home from work, school, or day care until at least 12-24 hours (US) and 24 hours (UK) after starting antibiotic treatment and some guidelines also recommend exclusion until resolution of symptoms [14-17]. Antibiotic chemoprophylaxis is also recommended for high-risk asymptomatic close contacts of iGAS cases [18]. Despite the impact of isolation from school or work, evidence for how long infectivity is likely to persist once treatment has commenced has not been reviewed systematically. We aimed to fill this evidence gap by reviewing studies which reported time from initiation of antibiotics to negative GAS culture in patients with confirmed pharyngeal GAS infection or carriage.

## METHODS

### Search strategy and selection criteria

Our protocol was registered with PROSPERO (CRD42021290364) on 8 November 2021. A professional librarian [MC] searched Ovid MEDLINE (1946-), EMBASE (1974-) and the Cochrane library on 18 October 2021 without language restrictions (see Supplementary Appendix A for search terms). Bibliographies of systematic reviews identified were searched for eligible studies. Studies included met the following criteria (see Supplementary Appendix B for full criteria): 1) peer-reviewed primary research with ten or more participants; 2) patients with GAS pharyngitis, scarlet fever or asymptomatic pharyngeal GAS carriage confirmed according to methods in ‘*Laboratory diagnosis* of group *A streptococcal infections’* or which reported diagnosis specifically of ‘group A’ streptococcal infection; 3) antibiotic administered to participants; 4) reported rates or counts of patients with positive or negative throat cultures for GAS whilst on antibiotic treatment, or time to clearance following initiation of antibiotics [19]. Two reviewers (EM, AL) independently screened titles and abstracts using the rayyan.ai platform [20]. Risk of bias was assessed independently and in parallel by two reviewers (EM, AL) using National Institutes of Health study quality assessment tools [21]. At all stages, disagreements were resolved by discussion between the two reviewers with arbitration if required by a third reviewer (CB, TL).

### Data analysis

Data from each study were extracted by one reviewer (EM) and cross-checked by a second reviewer (AL) into a custom Microsoft Excel form. The main data items extracted were: characteristics of study (design, location), patients (age, sex), treatment (drug, administration route, dose and duration), GAS identification method, and frequencies of study participants who were tested and who were culture-positive on day 1, day 2 and days 3-9 from start of antibiotic treatment. For the primary outcome of time to negative throat culture on antibiotic treatment, we conducted random effects meta-analysis to calculate pooled proportions of study participants who remained culture-positive at each time point. Where studies reported culture results at other time points, we mapped these to the later corresponding time-point. For example, a study reporting on days 1-2 would be included in the day 2 pooled estimate.

All statistical analyses were performed using STATA version 15.0 (StataCorp, College Station, Texas, USA). The score method was used to produce study specific confidence intervals (CIs) and the Freeman-Tukey double arcsine transformation was used to calculate weighted pooled proportion estimates for each sub-group [22]. The Wald method was used to produce CIs for these pooled estimates and heterogeneity within subgroups was tested using the *X*^2^ test and quantified by the *I*^*2*^ statistic. Meta-analysis was performed for all antibiotics combined and for subgroups by type of antibiotic (classes and individual types) using metaprop_one [23]. For the secondary outcome of culture positivity after completing antibiotic therapy, the same approach was used to calculate pooled proportions of participants who were culture-positive in the early (<72hrs), intermediate (72hrs-10 days), and late (>10 days) post-antibiotic period. Where studies reported typing or multilocus enzyme electrophoresis (MLEE), meta-analysis was performed using the same approach to calculate pooled proportions or participants who had documented clearance of GAS at the end of therapy followed by either relapse or reacquisition of the same strain of GAS or acquisition of a new strain of GAS after antibiotics, by class of antibiotic. The same approach was used to calculate pooled proportions reporting adverse drug reactions, by class of antibiotic. Meta-regression of proportion against day of culture (as ordinal variable) was performed using metapreg (to test for linear trend) and metareg (to generate bubble and line plots) [24, 25].

## RESULTS

Database searches identified 5,068 unique records of which 209 were selected for full text review. Five full texts could not be obtained, and 168 studies were excluded (Figure 1). Citation searches identified a further 26 studies, of which 25 were retrieved and 18 were excluded. Overall, 43 studies were included in the review representing observations on 7,168 patients with GAS confirmed on throat culture. Most were randomised controlled trials (n=37, 86%), three (7%) were non-randomised controlled trials, and three (7%) were observational ‘before and after’ studies (Table 1). A minority reported early culture results taken 24 hours (n=12, 28%) or 48 hours (n=9, 21%) after antibiotic initiation. Most reported outcomes only in children (n=29, 67%), 11 (26%) in children and adults, two in adults only, and the age range was not reported in one study. Pharyngitis (n=42, 98%) and tonsillitis (n=12, 28%) were the most studied clinical diagnoses, with few reporting on scarlet fever (n=5, 12%) or asymptomatic carriage (n=3, 7%). Most reported on the use of penicillins (n=34, 79%) or cephalosporins (n=14, 33%). Studies were predominantly conducted in North America (n=33, 77%) and Europe (n=6, 14%). Six (14%) studies were assessed as having a ‘low’ risk of bias, 20 (47%) ‘moderate’ risk of bias and 17 (40%) ‘high’ risk of bias (Supplementary Appendix C).

**Figure 1.**
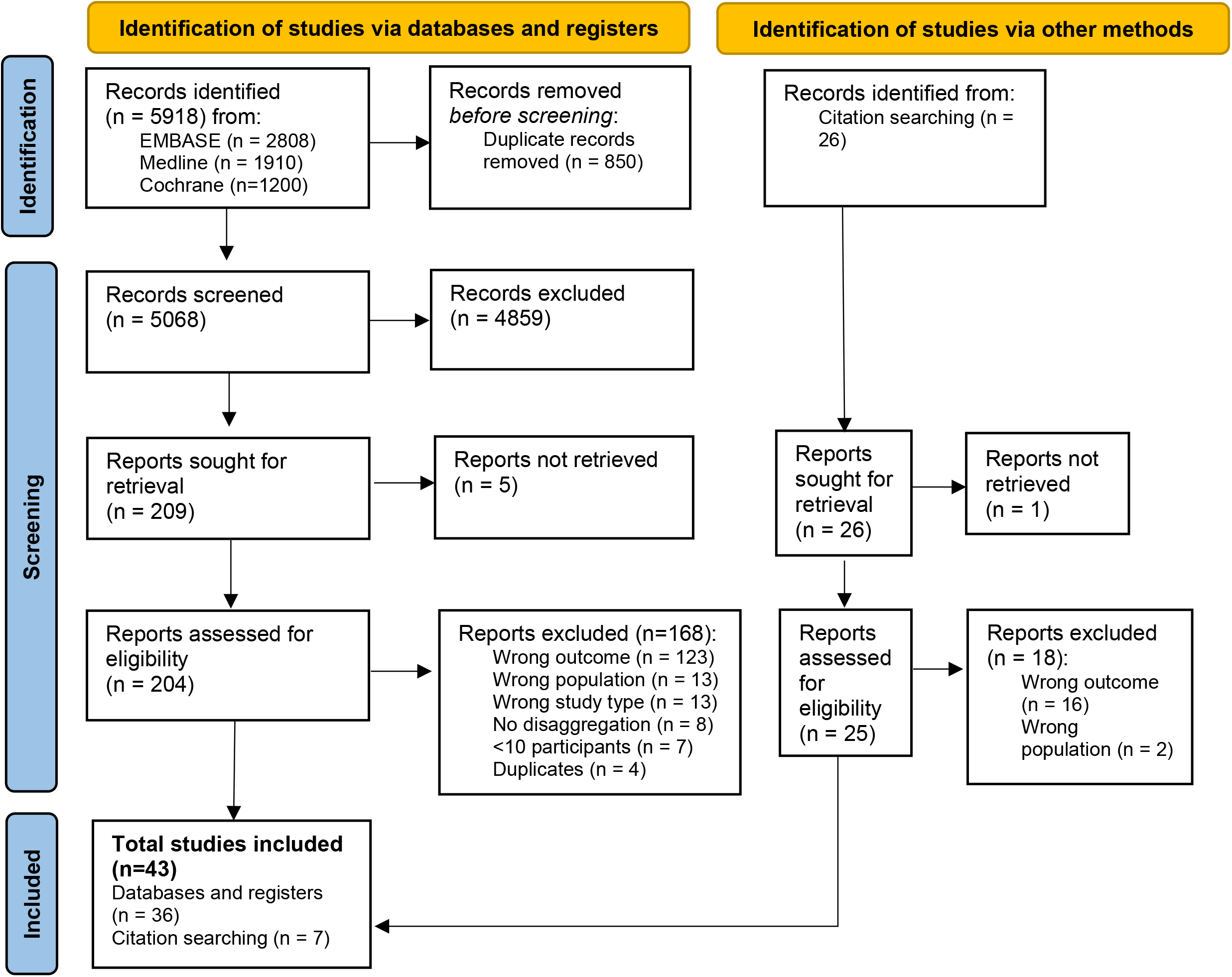
PRISMA flow diagram.

**Table 1.**
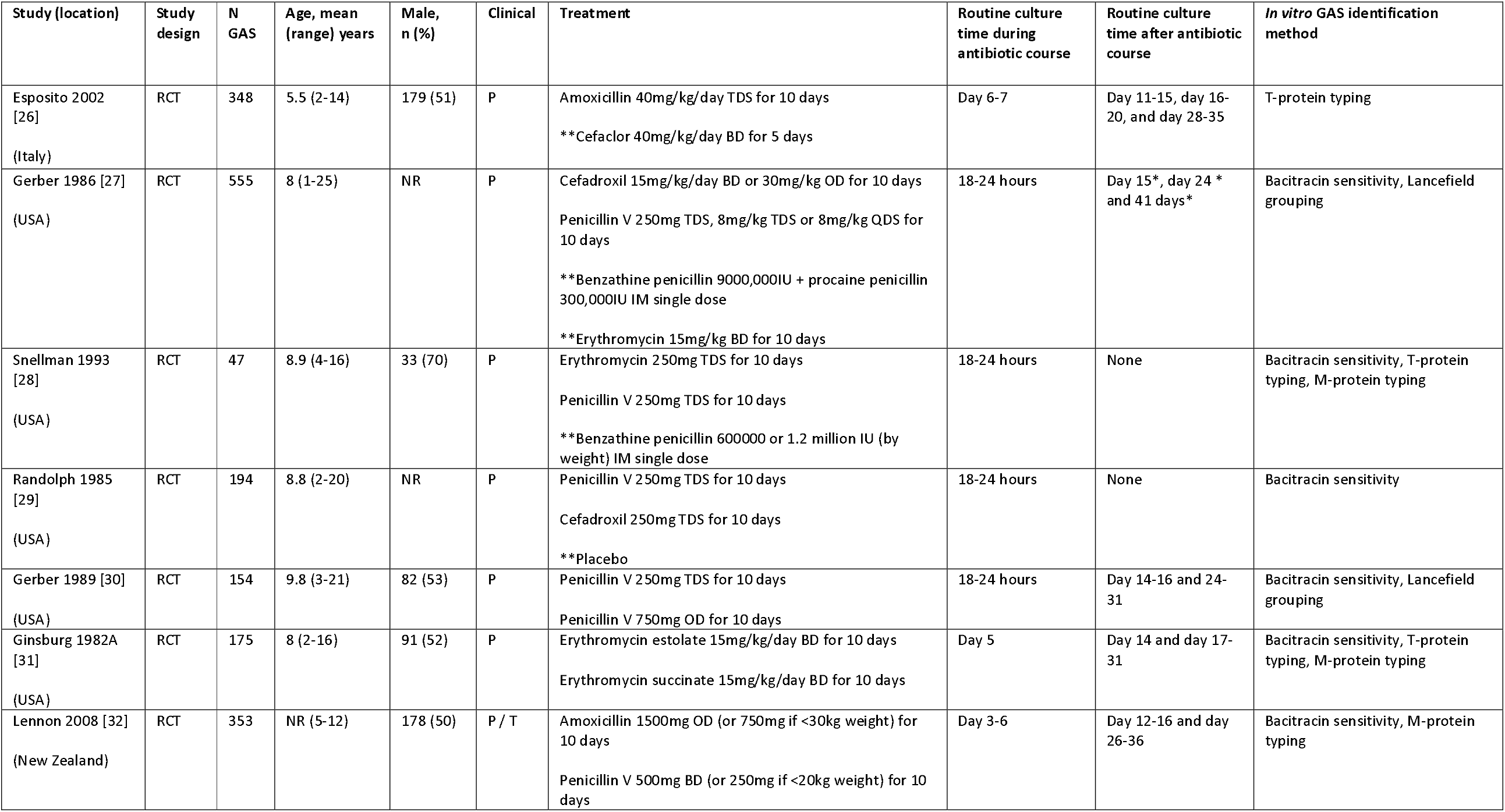
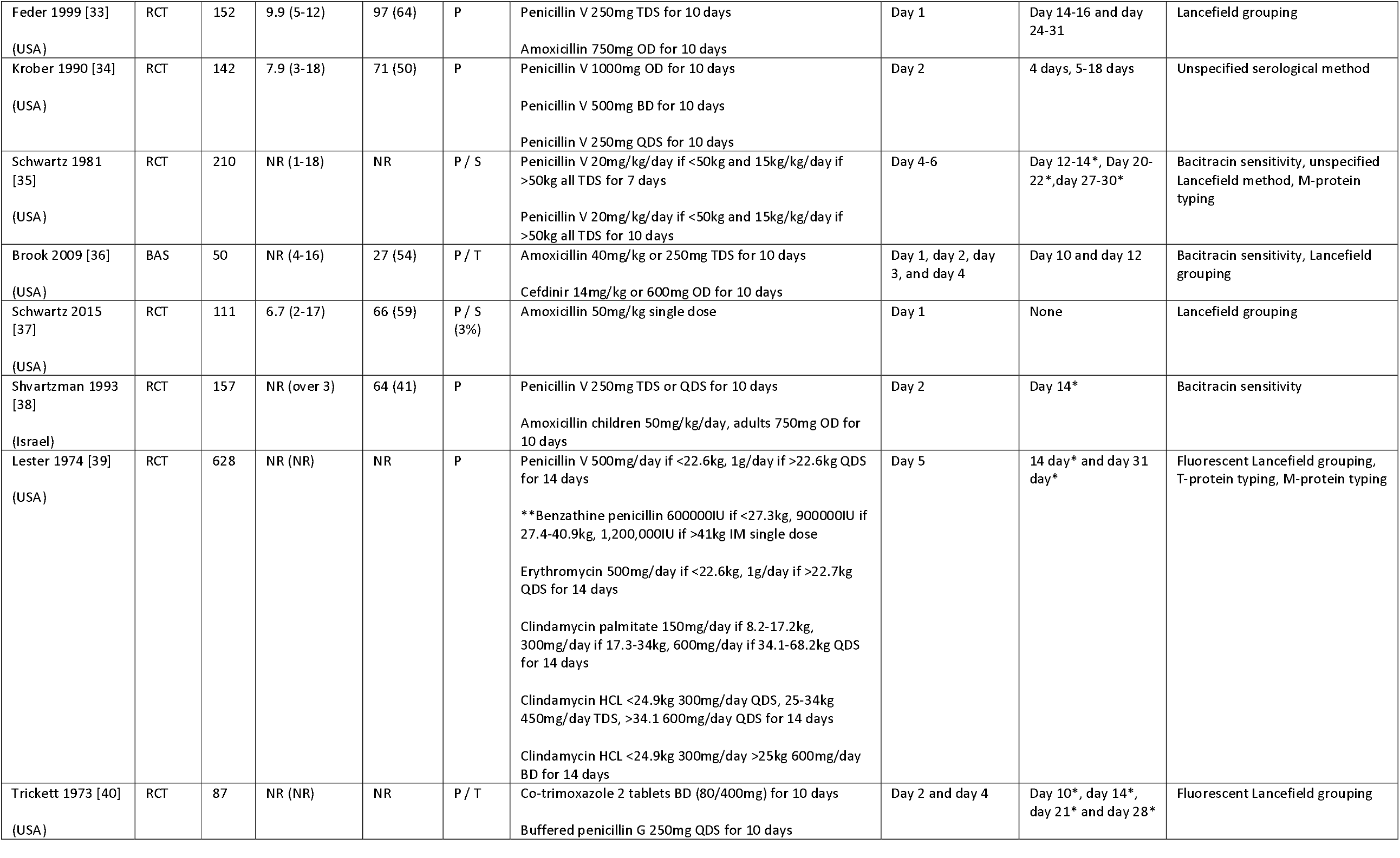
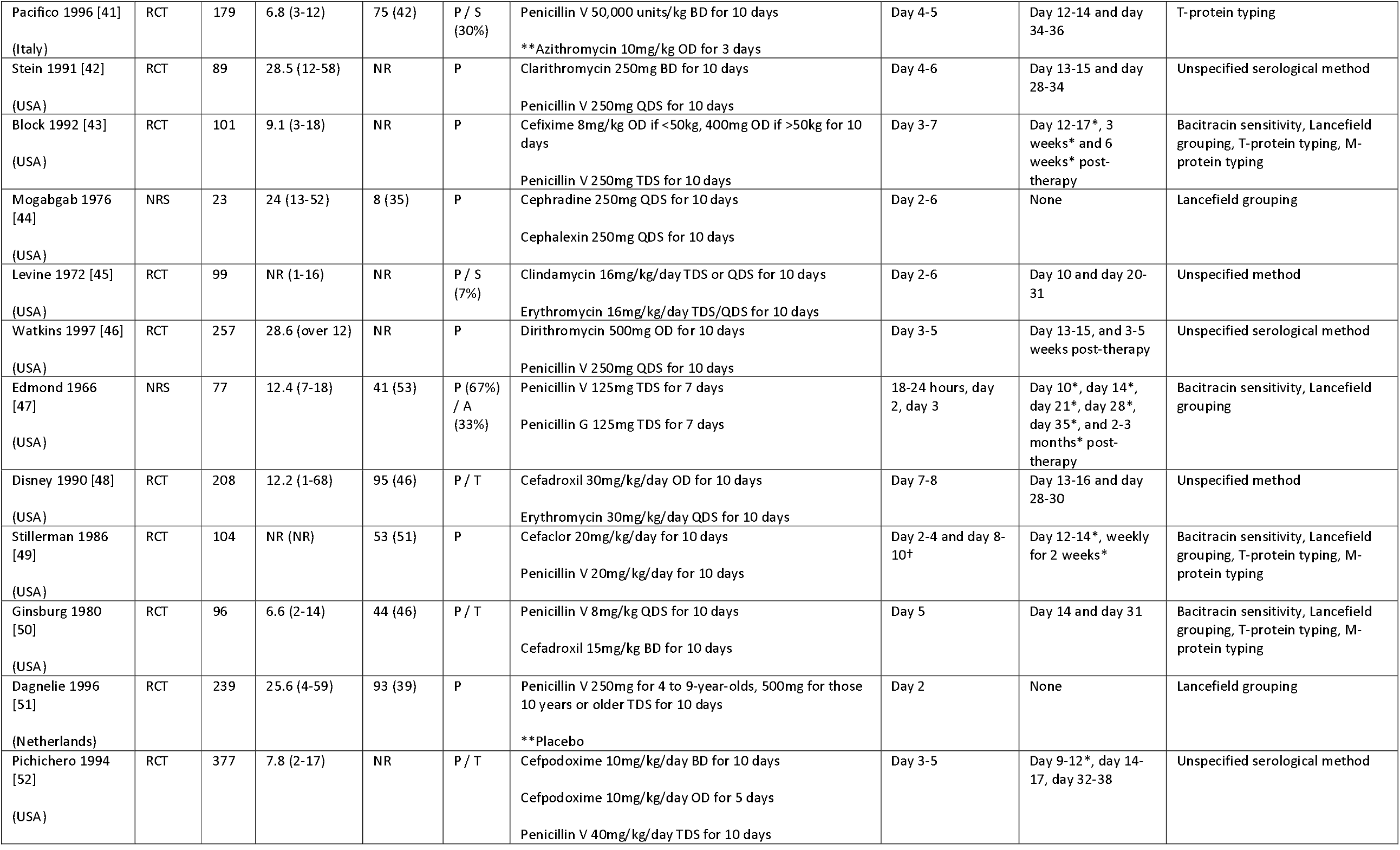
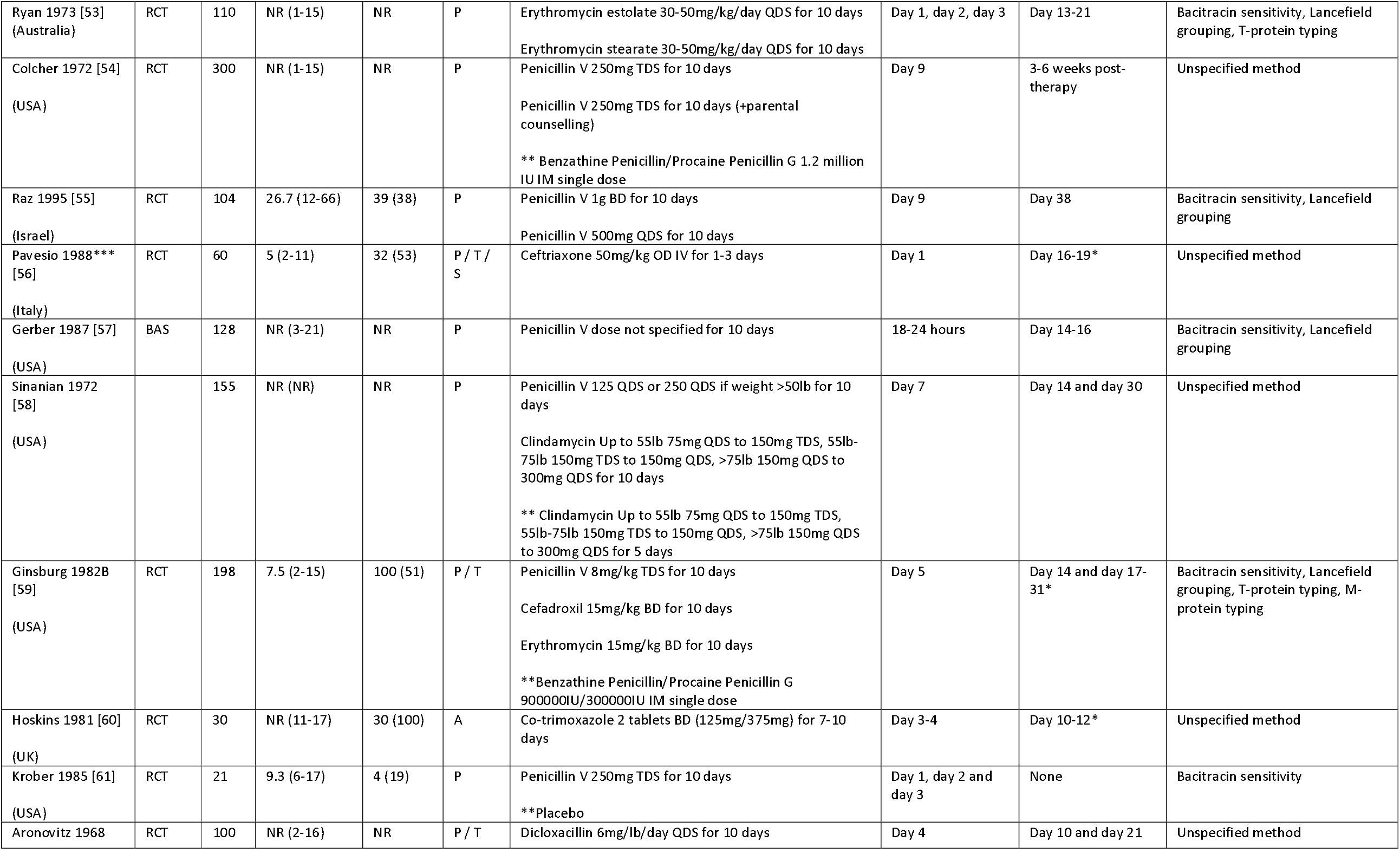
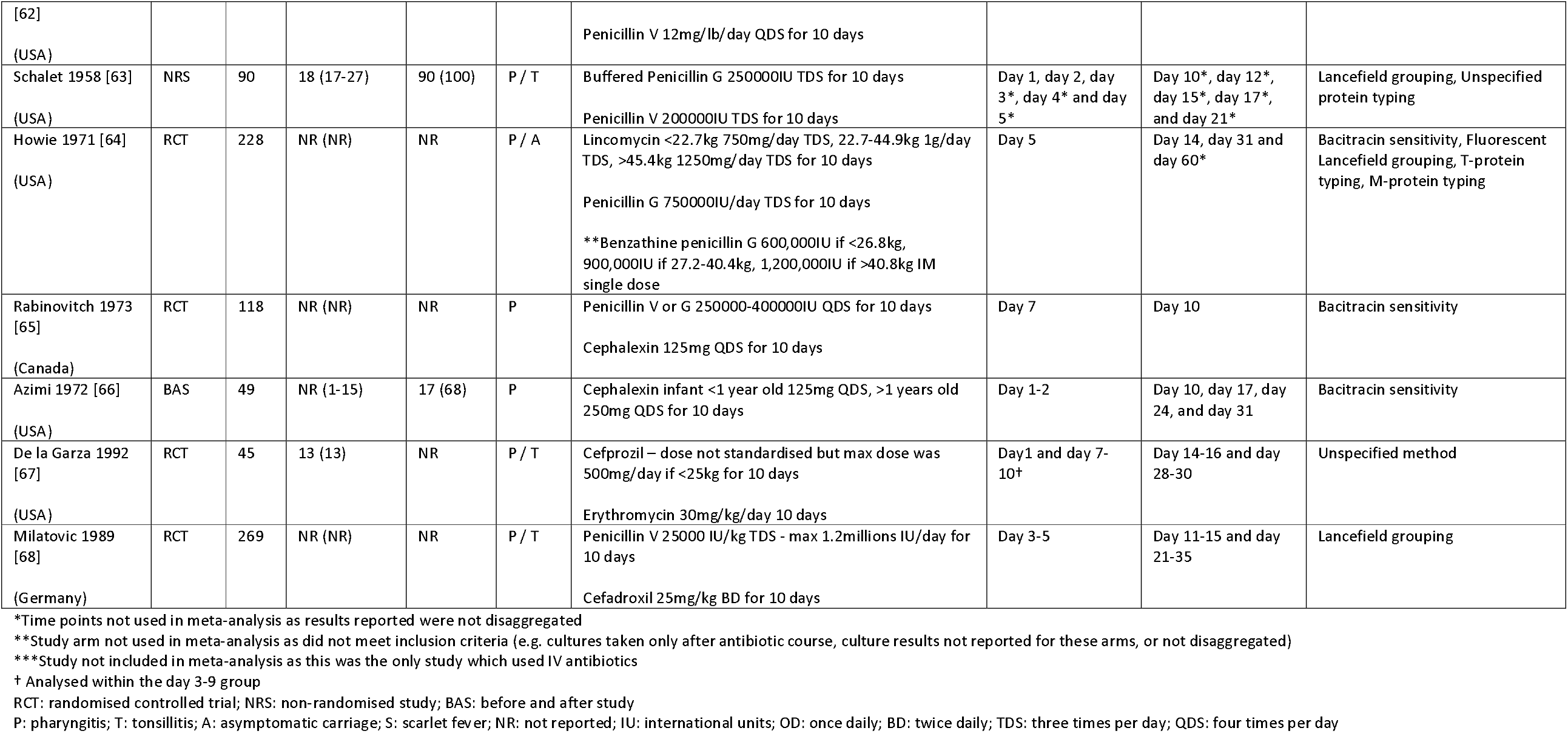
Characteristics of included studies (n=42)

Study methodologies were broadly similar; routine culture for beta-haemolytic streptococci was performed, most commonly on blood agar, with preliminary and confirmatory GAS identification. Throat swabs were repeated at pre-defined visits after initiation of treatment. Only reporting of bacterial culture for GAS was used to determine the primary and secondary outcomes in this review.

For all oral antibiotics combined (n=42 studies), the pooled proportion of individuals with confirmed GAS who remained culture-positive was 6.9% (95% CI 2.7-16.8%) on day 1, 5.4% (95% CI 2.1-13.3%) on day 2, and 2.6% (95% CI 1.6-4.2%) between days 3-9 (Figure 2). Brook *et al*. was an outlier, reporting high proportions of culture-positive patients at 24 hours (72.0%) and 48 hours (44.0%). Heterogeneity statistics and evidence of differences between sub-group pooled estimates are summarised in Supplementary Appendix E.

**Figure 2.**
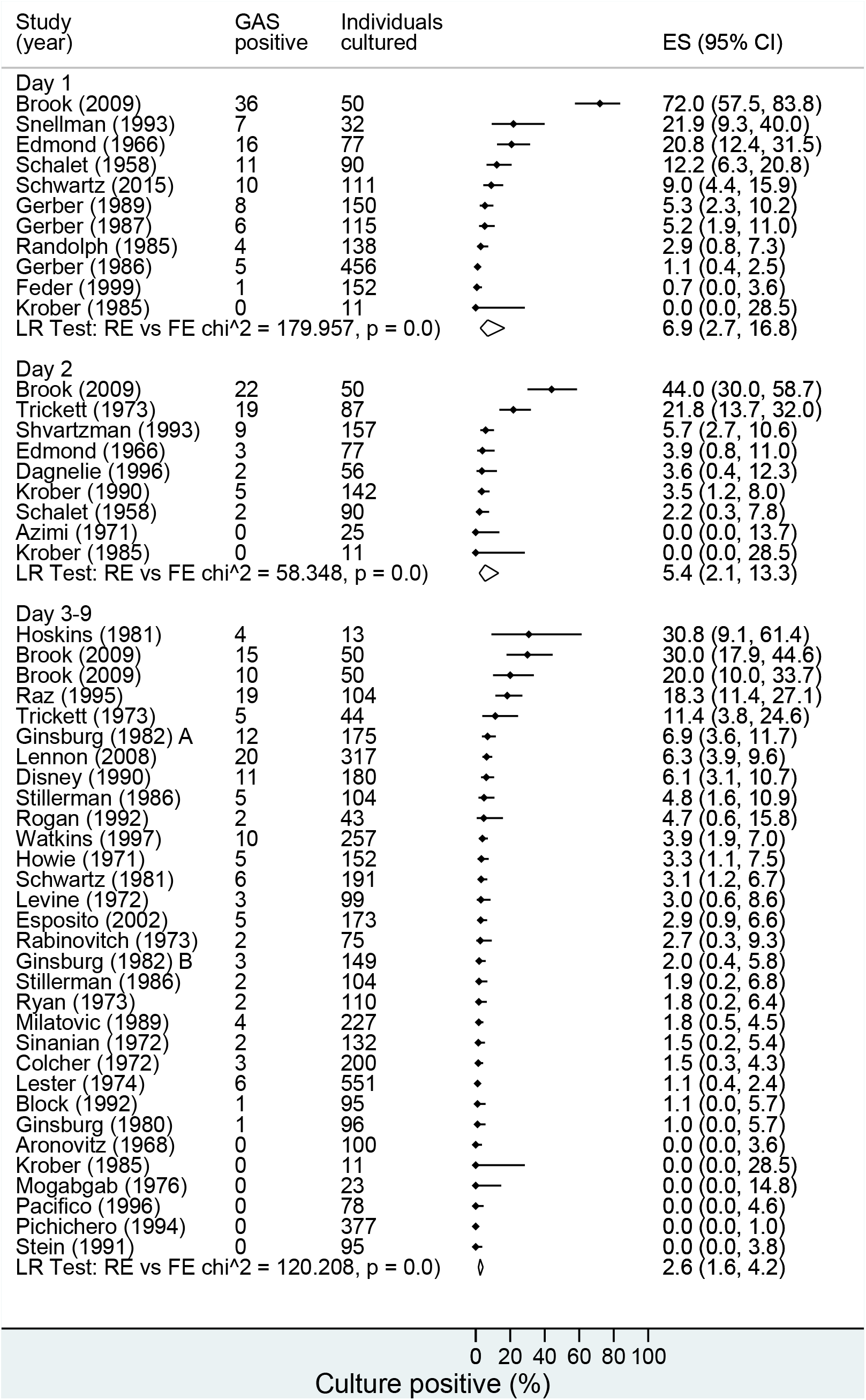
All antibiotic types - proportion of patients with culture-confirmed group A streptococcal (GAS) throat infection or carriage at day 1 (24 hours after the start of antibiotics), day 2 and days 3-9 (n=42 studies).

Antibiotic class subgroup analyses were possible for penicillins and cephalosporins. For penicillins (Figure 3), positivity was 6.5% (95% CI 2.5-16.1%) on day 1, 4.7% (95% CI 1.7-12.4) on day 2, and 2.6% (95% CI 1.4-4.8%) on days 3-9. For cephalosporins (Figure 4), positivity was 1.6% on day 1 (95% CI 0.04-42.9%), 16.0% (95% CI 8.2-28.9%) on day 2, and 0.8% (95%CI 0.2-3.5%) on days 3-9. Brook et al. was an outlier reporting high percentage positivity as previously noted.

**Figure 3.**
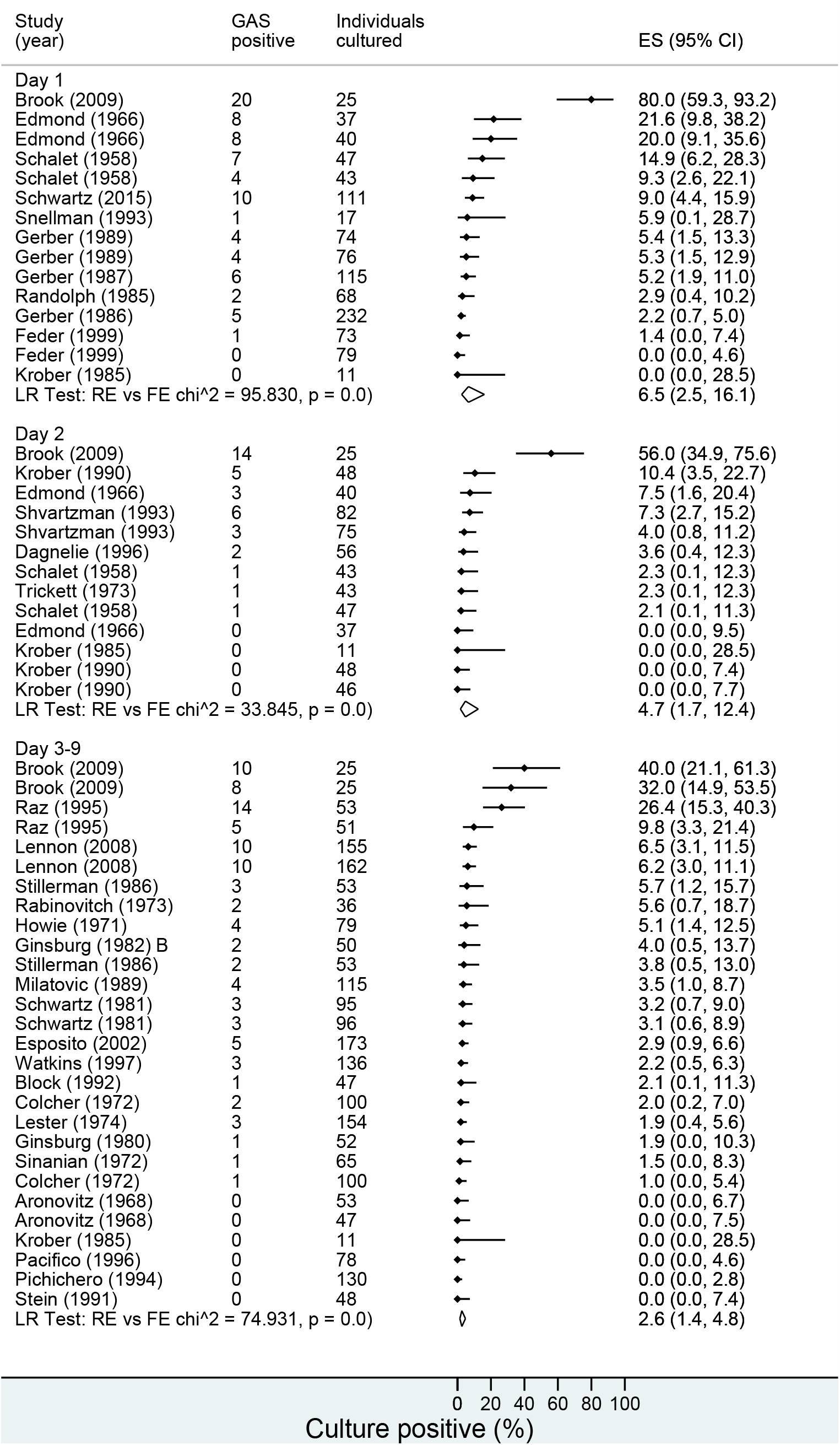
Penicillins - proportion of patients with culture-confirmed group A streptococcal (GAS) throat infection or carriage at day 1 (24 hours after the start of antibiotics), day 2 and days 3-9 (n=34 studies).

**Figure 4.**
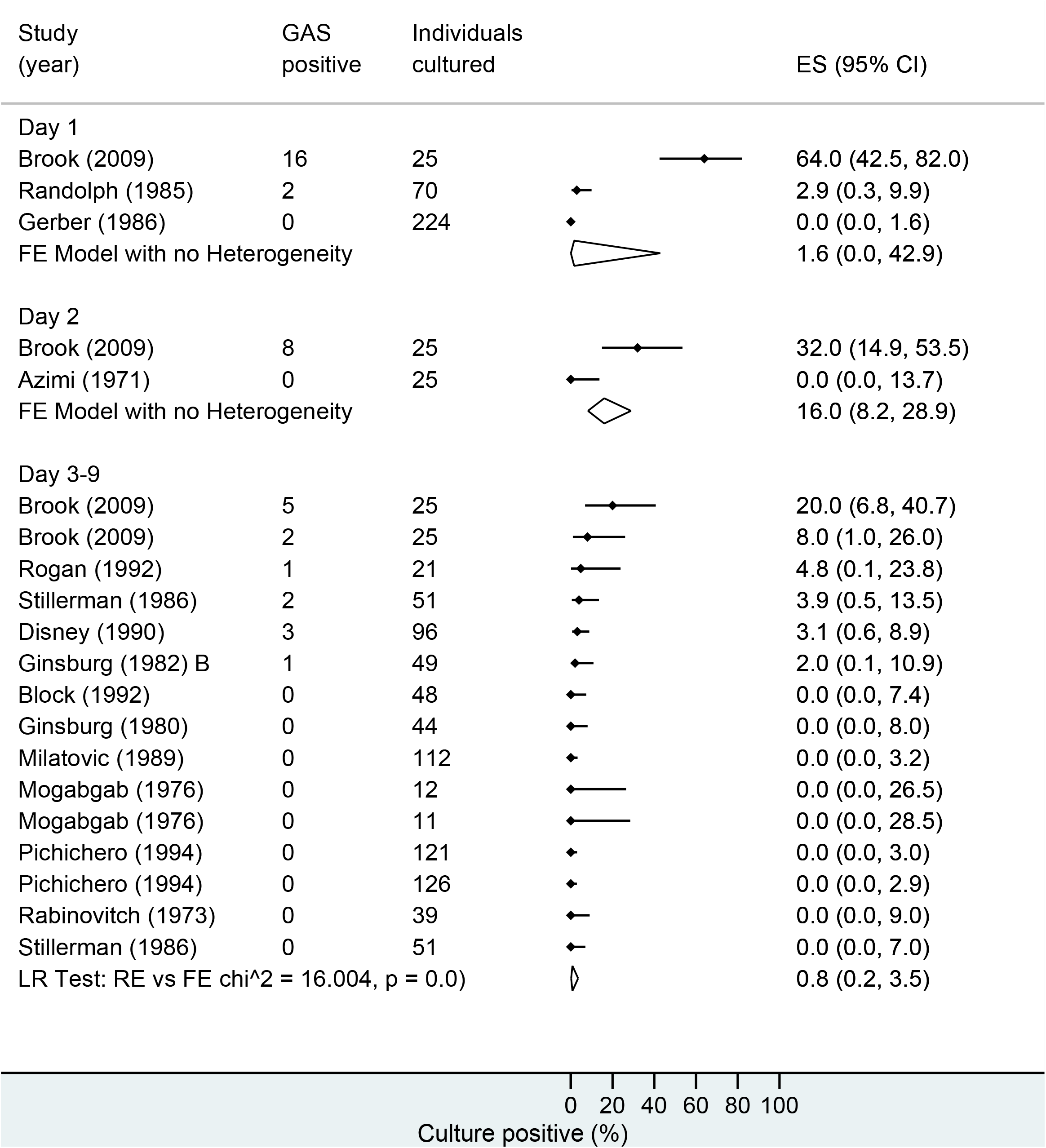
Cephalosporins - proportion of patients with culture-confirmed group A streptococcal (GAS) throat infection or carriage at day 1 (24 hours after the start of antibiotics), day 2 and days 3-9 (n=14 studies).

Meta-analysis by single antibiotic agent was possible only for penicillin V (Supplementary Figure S1). This showed day 1 positivity of 4.7% (95% CI 2.5-8.5%), day 2 positivity of 3.8% (95% CI 2.3-6.3), and day 3-9 positivity of 2.1% (95% CI 1.2-4.0%).

Meta-regression by day of culture did not demonstrate evidence for a linear effect of increasing time on proportion culture-positive for all antibiotics (p=0.12) (Supplementary Figure S2) nor trends for penicillins (p=0.21) (Supplementary Figure S3), cephalosporins (p=0.23) (Supplementary Figure S4) or Penicillin V (p=0.71) (Supplementary Figure S5).

Only 2 studies reported culture results within 24 hours of starting therapy. Snellman et al. reported on throat cultures taken on 3 occasions within the first 24 hours [28]. Of the patients who cleared GAS within 24 hours, the median time to clearance with any antibiotic (oral penicillin, intramuscular (IM) penicillin and oral erythromycin) was 18 hours. However, 8/47 (17.0%) patients remained GAS culture-positive at 24 hours and their results were not disaggregated by antibiotic. Schwartz *et al*. reported throat culture results 11-24 hours after a single dose of amoxicillin; 10/111 (9.0%) remained culture-positive [37].

Ten studies investigated macrolides [28, 31, 39, 42, 45, 46, 48, 53, 59, 67], three clindamycin [39, 45, 58] and two co-trimoxazole [40, 60] with a wide range of dosing regimens (Supplementary Appendix D). Only one study reported on throat cultures taken at 24 hours following initiation of treatment with a macrolide; 6/15 (40%) remained culture positive [28]. However, another study reported that 2/110 (1.8%) were culture positive on 1-3 days after treatment initiation [53]. Regarding lincosamides, we did not identify any studies reporting on cultures taken within 48 hours of treatment initiation, however, results from the three studies reporting on later time periods were in keeping with our overall findings. Two studies reporting on the use of sulphonamides found comparatively high rates of culture-positivity; 18/44 (40.9%) and 4/13 (30.8%) remained culture positive on day 2 and days 3-4 following initiation of co-trimoxazole, respectively [40, 60].

A wide range of oral penicillin V regimens was used; frequencies ranged from one to four times daily, with a variety of weight or age-based doses in children and adult doses ranging from 500mg to 2g per day. Only three studies directly compared oral penicillin V dosing regimens [30, 34, 55]. Gerber et al. compared 750mg once daily to 250mg three times daily penicillin V, reporting that after 18-24 hours 4/76 (5.3%) patients vs. 4/74 (5.4%) patients were culture-positive, respectively. Krober *et al*. compared three dosing regimens of penicillin V; 1000mg once daily vs 500mg twice daily vs 250mg four times daily. After two days of therapy 5/48 (10.4%), 0/48 (0%) and 0/46 (0%) had positive throat cultures, respectively. Raz *et al*. compared penicillin V 1g twice daily to 500mg four times daily and reported that cultures taken after 9 days of treatment were positive in 5/51 (9.8%) patients and 14/53 (26.4%) patients, respectively. Similarly, eight cephalosporins were investigated, from first to third generation. There were insufficient results for each regimen to support subgroup meta-analyses by penicillin dosing strategy or cephalosporin type.

Several studies compared the efficacy of once daily amoxicillin against penicillin V with broadly similar rates of culture positivity at equivalent time points [32, 33, 38]. One compared penicillin V 250mg three times daily to amoxicillin 750mg once daily, with 1/73 (1.4%) and 0/79 (0.0%) culture-positive after 18-24 hours, respectively [33]. Another compared penicillin V taken 3 or 4 times daily to daily amoxicillin, reporting culture-positivity on day 1 of 26/82 (7.3%) for penicillin and 3/75 (4%) for amoxicillin [38]. A third study compared 500mg penicillin V twice daily and once daily amoxicillin, with 10/162 (6.2%) and 10/155 (6.5%) patients found to be culture positive after 3-6 days of antibiotics, respectively [32].

Three studies used placebo control arms [29, 51, 61]. Dagnelie *et al*. reported 2/56 (3.6%) patients taking oral penicillin V remained culture-positive after 48 hours, compared to 41/55 (74.5%) of those taking placebo [51]. Similarly, Randolph *et al*. found that 100% of patients with GAS pharyngitis who received placebo remained positive at 18-24 hours, compared to ‘approximately 3%’ of those in the oral penicillin and cefadroxil groups [29].

Five studies reported IM penicillin therapy; results were consistent with our findings for antibiotics overall [28, 39, 54, 59, 64]. Snellman *et al*. found 1/15 (6.7%) children were culture-positive at 24 hours following single dose benzathine penicillin [28]. Lester *et al*. reported no positive cases at day 5 among 126 patients treated either with Benzathine penicillin or combined Benzathine Penicillin/Procaine Penicillin G [39]. Colcher *et al*. found 3/100 (3%) to be culture-positive at day 9 following a single dose of Benzathine Penicillin/Procaine Penicillin G [54].

Only one study was identified that reported the effect of an intravenous (IV) antibiotic [56]. This study investigated the treatment of GAS tonsillitis or scarlet fever in 60 children who received either a single dose or 3-days’ IV ceftriaxone 50mg/kg dose. All patients in both arms were culture negative 24 hours after the first dose of IV ceftriaxone.

In studies in which throat cultures were taken at routine times after completion of antibiotics (n=23), the proportion of patients who remained culture-positive was 9.1% (95% CI 7.3-11.3) overall; 3.1% (95% CI 1.0-9.5) within 72 hours, 8.9% (95% CI 6.8-11.6) at 72 hours to 10 days after completion, and 9.9% (95% CI 7.8-12.7) at 10 or more days (Supplementary Figure S6). Thirteen studies compared the GAS strain identified after completion of antibiotics and initial culture clearance to the original strain; eight by M typing, one by T typing, one by MLEE and three by an unspecified serotyping method. Among these studies the overall pooled proportion that had documented culture clearance after therapy followed by a further positive GAS throat culture was 13.3% (95% CI 10.5-16.7); 10.4% (95% CI 8.6-12.5) had relapse or reacquisition of the original GAS strain and 2.7% (95% CI 1.7-4.3) had acquired a new strain of GAS after therapy (Figures 5, and Supplementary Figures S7 and S8).

**Figure 5.**
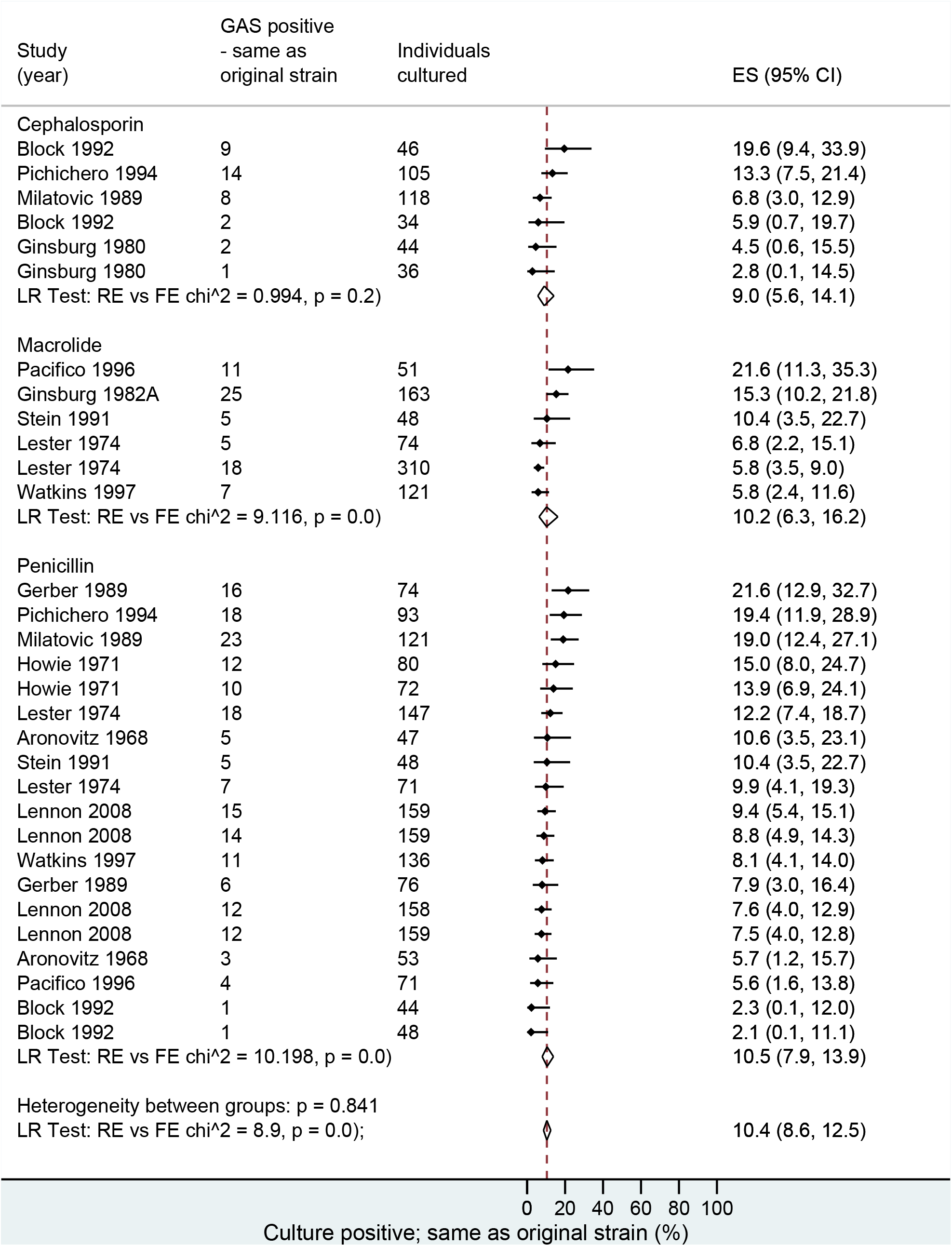
Proportion of patients with culture-confirmed group A streptococcal (GAS) throat carriage after completion of antibiotics (including relapse/reacquisition of the same strain and acquisition of a new strain) by antibiotic class, where typing was reported (n=13 studies).

There were insufficient observations to support sub-group meta-analysis by clinical indication (tonsillitis, pharyngitis, scarlet fever or asymptomatic carriage). Five studies included individuals with scarlet fever, but none provided disaggregated results for this sub-group [35, 37, 41, 45, 56]. Three studies included individuals with asymptomatic carriage, [47, 60, 64] but only Hoskins *et al*. reported distinct results for this sub-group. This was a small paediatric study in which throat cultures were repeated 3-4 days after starting co-trimoxazole, at which point 4/13 (30.8%) children remained culture positive. As only two studies reported exclusively on adults, meta-analysis by age group was not possible [40, 63]. No studies were identified that specifically reported on patients at risk of severe GAS disease, such as pregnant women, the elderly or immunocompromised individuals.

Where reported, the proportion of patients who reported a side effect or adverse drug reaction was 6.0% (95% CI 2.4-14.3) (n=14 studies) and the proportion who ceased the study drug due to an adverse reaction was 0.3% (95% CI 0.1-1.7) (n=12 studies) (Supplementary Figures S9 and S10).

## DISCUSSION

The key finding of our systematic review and meta-analysis is that antibiotic treatment achieves a high rate (>90%) of clearance of pharyngeal GAS 24 hours after initiation of therapy. This evidence supports the public health guidelines that recommend that individuals with GAS pharyngitis or scarlet fever isolate for at least 24 hours after starting antibiotic treatment, given the high secondary attack rate for GAS and the need to prevent transmission. There was some evidence of a waning effect following completion of antibiotic treatment; GAS was cultured from the pharynx of almost 10% of patients on routine follow-up 10 or more days after completion of antibiotics and where typing was performed. The majority were confirmed to be relapse or reacquisition of the original GAS strain. This supports the need for clearance throat swabs for individuals potentially acting as a reservoir of infection in iGAS outbreaks. We mitigated for patients with symptoms being more likely to return for follow-up as a source of bias by only including estimates where all participants were scheduled for repeat specimens, regardless of symptoms.

Between-study heterogeneity gave rise to overlapping confidence intervals for sub-group estimates at each time point, and meta-regression did not provide evidence that culture-positivity was higher on day 1 than on day 2 or days 3-9 (assuming a linear effect). Our analysis was also unable to discern differences between antibiotic classes or types or between different dosing regimens for the same antibiotic. The primary focus of this study was clearance of pharyngeal GAS during antibiotic treatment and immediately after treatment completion; therefore, the optimal duration of antibiotics for longer term eradication was outside of our scope. However, several reviews have examined this issue [69-72]. Casey *et al*. and Holm *et al*. found that short course (<5 days) penicillin V was inferior and cephalosporins were superior in terms of bacterial eradication when compared to 10 days of penicillin V. Altamimi et al. defined more specific outcomes of early and late bacteriological failure as isolation of the same strain of GAS within two weeks and more than 2 weeks after completing therapy, respectively. Most included studies reported on macrolides, however some reported on penicillins or cephalosporins. They found that short (2-6 days) therapy was equivalent to standard (10 days) penicillin V for both outcomes when studies with low dose azithromycin were excluded.

A limitation of our review is that most included studies had a moderate or high risk of bias. There were several outliers among estimates from included studies, notably Brook *et al*. who conducted a before-and-after study comparing the proportion with positive culture among 50 children with GAS pharyngitis treated with either amoxicillin or cefdinir [36]. While the 40mg/kg dose of amoxicillin used in this study is lower than the 50mg/kg recommended in national guidance in the USA, this was the same dose used in 2 other included studies which found much lower rates of culture positivity at comparable timepoints [26, 33, 73]. This suggests that methodology rather than underdosing might explain their outlying findings. The Brook *et al*. study was assessed as having a moderate risk of bias because outcome assessors were not blinded and the proportion of eligible patients who were enrolled was unclear. However, other studies had a similar risk of bias; therefore, the reason for their particularly high percentages is unknown. Our findings might be affected by antibiotic carry-over, when a high concentration of antibiotic in the sample inhibits *in vitro* growth [74]. This effect would lower the culture-positive proportion in samples taken during or shortly after the antibiotic course.

The appropriate dosing of penicillin V for GAS pharyngitis is unclear. Debate surrounds the efficacy of twice daily dosing, and more frequent dosing intervals have been associated with reduced adherence [75, 76]. The two identified studies in which penicillin V dosing regimens were directly compared suggested that less frequent dosing delivered similar clearance rates, although Gerber *et al*. considered once daily penicillin V to be inferior to multiple daily dosing with regards to re-acquisition of infection following treatment [30]. Treatment with once daily amoxicillin also appeared to have the same effectiveness as standard multi-dosed penicillin V.

Most patients included in this review were treated with penicillins or cephalosporins, and there remains uncertainty regarding the performance of non-beta-lactam antibiotics, particularly in the early stages of treatment. Unfortunately, studies reporting on macrolides were limited and the only study to report on day 1 culture conversion included only 15 participants in the macrolide arm [28]. Two meta-analyses evaluated clinical and bacteriological outcomes with clarithromycin, both reporting favourable bacteriological clearance compared with other antibiotics [77, 78]. However, these reviews assessed bacterial eradication after the antibiotic course.

GAS has retained exquisite *in vitro* sensitivity to penicillin. However, while the prevalence of *erm* genes conferring resistance to macrolides and lincosamides in GAS bacteraemia in England has remained steady at 8% and 9% respectively, an increasing prevalence of both erythromycin and clindamycin resistance has been reported in iGAS cases in USA, reaching 22.8% and 22.0% respectively in 2017 [1, 79]. Studies investigating macrolide treatments have explicitly excluded patients with resistant organisms from culture endpoint analysis. Therefore, the prevalence and presence of macrolide resistance is an important consideration when determining optimal treatments. Clindamycin has demonstrated inhibition of GAS virulence factors *in vivo* and has been recommended as part of combination therapy in severe forms of GAS infection such as necrotising fasciitis and streptococcal toxic shock syndrome for anti-toxin ribosomal effects [80, 81]. However, its use in eradicating GAS from the pharynx must be balanced against tolerability considerations. The two studies in our review that investigated co-trimoxazole demonstrated relatively low rates of early culture conversion suggesting this is a sub-optimal antibiotic for prevention of GAS transmission in the context of GAS outbreaks.

Although rates of reported penicillin allergy are approximately 10%, the prevalence of true penicillin allergy is much lower and rates of anaphylactic penicillin allergy in the UK and USA are <1% [82, 83]. Paucity of evidence regarding optimal alternatives such as macrolides and lincosamides requires urgent attention, particularly as resistance varies by region. Evidence to support antibiotic choice for GAS patients with penicillin allergy in settings where macrolide resistance was suspected or present is limited [84]. There were no studies investigating GAS clearance by alternative non-beta-lactam agents such as oxolidazones, fluoroquinolones, tetracyclines, rifampicin, tetracycline, chloramphenicol, glycopeptides or lipoglycopeptides. Antibiotics were generally well tolerated regardless of class. Although rates of reported side effects were higher in non-beta-lactam antibiotics, cessation of the study drug due to side effects was rare for any class of antibiotic.

Presence of symptoms could hypothetically be correlated with culture clearance because inflammation can influence tissue perfusion and antibiotic concentrations. However, only one study investigated effectiveness of clearing asymptomatic GAS carriage and no firm conclusions could be drawn as this was a small study with a sub-optimal antibiotic [60]. This highlights an important evidence gap relating to the use of antibiotics to decolonise asymptomatic contacts identified as potential reservoirs of GAS transmission in outbreak situations. Similarly, the evidence base for optimal treatment of patients with scarlet fever is limited.

Only studies reporting on pharyngeal GAS were reviewed and our results cannot be generalised to infections at other sites. We excluded studies examining the efficacy of antibiotics in individuals with recurrent GAS pharyngitis or persistent colonisation despite initial treatment. Factors contributing to antibiotic failure in the absence of antimicrobial resistance are poorly understood, but co-pathogenicity with beta-lactamase-producing flora, lack of compliance, the carrier state, poor antibiotic penetration into tonsillar tissues and re-infections may contribute.[85, 86] Our post-antibiotic results are in keeping with other meta-analyses that have looked at bacteriological eradication after therapy for GAS pharyngotonsillitis [87]. A minority of studies in our review performed typing to distinguish between relapse and reacquisition with the original GAS strain and acquisition of a GAS new strain, however, the latter appears to be far less common. This may be an important consideration during prolonged outbreaks.

Whilst most studies in our review reported use of standard bacitracin sensitivity and Lancefield grouping, the identification method was not reported in all studies. Furthermore, some studies used uncommon typing methods. Molecular diagnostic techniques are increasingly used in clinical microbiology and, whilst these approaches are more sensitive, it is not known whether a positive molecular diagnostic result carries the same public health risk as culture. Future studies need to examine positivity based on molecular, point-of-care and culture testing. Further research is needed to quantify clearance rates at distinct periods within the first 24 hours of treatment and to determine whether non-beta-lactam antibiotics are effective, particularly when macrolide resistance is present or suspected and in patients who are asymptomatic or at high risk of severe GAS infection. Our review has generated evidence supporting a time interval of 24 hours for isolation for work or school following initiation of treatment for pharyngeal group A streptococcal infection.

## Data Availability

Participant identifiable data were not collected or held for this study. Data collected for the study may be made available on request.

## REFERENCES

1. UKHSA. Laboratory surveillance of pyogenic and non-pyogenic streptococcal bacteraemia in England: 2020 update. UK Health Security Agency; 2021 23 November 2021. Contract No.: 19.

2. Oliver J, Malliya Wadu E, Pierse N, Moreland NJ, Williamson DA, Baker MG. Group A Streptococcus pharyngitis and pharyngeal carriage: A meta-analysis. PLoS Negl Trop Dis. 2018;12(3):e0006335.

3. Pearson M, Fallowfield JL, Davey T, Thorpe NM, Allsopp AJ, Shaw A, et al. Asymptomatic group A Streptococcal throat carriage in Royal Marines recruits and Young Officers. J Infect. 2017;74(6):585–9.

4. Spitzer J, Hennessy E, Neville L. High group A streptococcal carriage in the Orthodox Jewish community of north Hackney. Br J Gen Pract. 2001;51(463):101–5.

5. Cordery R PA, Begum L, Mills E, Mosavie M, Vieira A, Jauneikaite E, Leung RCY, Siggins MK, Ready D, Hoffman P, Lamagni T, Sriskandan S.. Frequency of transmission, asymptomatic shedding, and airborne spread of Streptococcus pyogenes among schoolchildren exposed to scarlet fever: a longitudinal multi-cohort molecular epidemiology contact tracing study. 2021.

6. Daneman N, Green KA, Low DE, Simor AE, Willey B, Schwartz B, et al. Surveillance for hospital outbreaks of invasive group a streptococcal infections in Ontario, Canada, 1992 to 2000. Ann Intern Med. 2007;147(4):234–41.

7. Daneman N, McGeer A, Low DE, Tyrrell G, Simor AE, McArthur M, et al. Hospital-acquired invasive group a streptococcal infections in Ontario, Canada, 1992-2000. Clin Infect Dis. 2005;41(3):334–42.

8. Breese BB, Disney FA. Factors influencing the spread of beta hemolytic streptococcal infections within the family group. Pediatrics. 1956;17(6):834–8.

9. Falck G, Kjellander J. Outbreak of group A streptococcal infection in a day-care center. Pediatr Infect Dis J. 1992;11(11):914–9.

10. Feeney KT, Dowse GK, Keil AD, Mackaay C, McLellan D. Epidemiological features and control of an outbreak of scarlet fever in a Perth primary school. Commun Dis Intell Q Rep. 2005;29(4):386–90.

11. Adebanjo T, Apostol M, Alden N, Petit S, Tunali A, Torres S, et al. Evaluating Household Transmission of Invasive Group A Streptococcus Disease in the United States Using Population-based Surveillance Data, 2013-2016. Clin Infect Dis. 2020;70(7):1478–81.

12. Gerber MA, Baltimore RS, Eaton CB, Gewitz M, Rowley AH, Shulman ST, et al. Prevention of rheumatic fever and diagnosis and treatment of acute Streptococcal pharyngitis: a scientific statement from the American Heart Association Rheumatic Fever, Endocarditis, and Kawasaki Disease Committee of the Council on Cardiovascular Disease in the Young, the Interdisciplinary Council on Functional Genomics and Translational Biology, and the Interdisciplinary Council on Quality of Care and Outcomes Research: endorsed by the American Academy of Pediatrics. Circulation. 2009;119(11):1541–51.

13. Spinks A, Glasziou PP, Del Mar CB. Antibiotics for sore throat. Cochrane Database Syst Rev. 2013(11):CD000023.

14. David W. Kimberlin EDB, Ruth Lynfield, Mark H. Sawyer. Group A Streptococcal Infections. Red Book: 2021–2024 Report of the Committee on Infectious Diseases, Committee on Infectious Diseases, American Academy of Pediatrics. 32 ed. USA 2021.

15. CDC. Group A Streptococcus Infections. Infection Control in Healthcare Personnel: Epidemiology and Control of Selected Infections Transmitted Among Healthcare Personnel and Patients In: Prevention CfDCa, editor. USA: Centres for Disease Control and Prevention; 2021.

16. Steer JA, Lamagni T, Healy B, Morgan M, Dryden M, Rao B, et al. Guidelines for prevention and control of group A streptococcal infection in acute healthcare and maternity settings in the UK. J Infect. 2012;64(1):1–18.

17. PHE. Guidelines for the public health management of scarlet fever outbreaks in schools, nurseries and other childcare settings. Public Health England; 2017.

18. Health Protection Agency GASWG. Interim UK guidelines for management of close community contacts of invasive group A streptococcal disease. Commun Dis Public Health. 2004;7(4):354–61.

19. Johnson DR KE, Sramek J, Bicova R, Havlicek J, Havlickova H, Motlova J, Kriz P; for World Health Organisation Geneva. Laboratory diagnosis of group A streptococcal infections. 1996.

20. Ouzzani M, Hammady H, Fedorowicz Z, Elmagarmid A. Rayyan-a web and mobile app for systematic reviews. Syst Rev. 2016;5(1):210.

21. Health NIo. Study quality assessment tools.. Bethesda: National Institutes of Health; 2020. 22.

22. Freeman MF TJ. Transformations Related to the Angular and the Square Root.. The Annals of Mathematical Statistics. 1950;21(4):607–711.

23. Nyaga VN, Arbyn M, Aerts M. Metaprop: a Stata command to perform meta-analysis of binomial data. Arch Public Health. 2014;72(1):39.

24. Harbord RM HJ. Meta-regression in Stata. Stata Journal 008;8:493–519.

25. Nyaga V. METAPREG: Stata module to compute fixed and random effects meta-analysis and meta-regression of proportions. Boston College Department of Economics; 2019.

26. Esposito S, Marchisio P, Bosis S, Droghetti R, Mattina R, Principi N. Comparative efficacy and safety of 5-day cefaclor and 10-day amoxycillin treatment of group A streptococcal pharyngitis in children. International journal of antimicrobial agents. 2002;20(1):28–33.

27. Gerber MA. A comparison of cefadroxil and penicillin V in the treatment of streptococcal pharyngitis in children. Drugs. 1986;32:29–32.

28. Snellman LW, Stang HJ, Stang JM, Johnson DR, Kaplan EL. Duration of positive throat cultures for group A streptococci after initiation of antibiotic therapy. Pediatrics. 1993;91(6):1166–70.

29. Randolph MF, Gerber MA, DeMeo KK, Wright L. Effect of antibiotic therapy on the clinical course of streptococcal pharyngitis. J Pediatr. 1985;106(6):870–5.

30. Gerber MA, olph MF, DeMeo K, Feder HM, Jr., Kaplan EL. Failure of once-daily penicillin V therapy for streptococcal pharyngitis. American Journal of Diseases of Children. 1989;143(2):153–5.

31. Ginsburg CM, McCracken GH, Jr., Steinberg JB, Crow SD, Dildy BF, Lancaster K, et al. Management of group A streptococcal pharyngitis: a randomized controlled study of twice-daily erythromycin ethylsuccinate versus erythromycin estolate. Pediatric Infectious Disease. 1982;1(6):384–7.

32. Lennon DR, Farrell E, Martin DR, Stewart JM. Once-daily amoxicillin versus twice-daily penicillin V in group A beta-haemolytic streptococcal pharyngitis. Archives of Disease in Childhood. 2008;93(6):474–8.

33. Feder HM, Jr., Gerber MA, olph MF, Stelmach PS, Kaplan EL. Once-daily therapy for streptococcal pharyngitis with amoxicillin. Pediatrics. 1999;103(1):47–51.

34. Krober MS, Weir MR, Themelis NJ, van Hamont JE. Optimal dosing interval for penicillin treatment of streptococcal pharyngitis. Clinical Pediatrics. 1990;29(11):646–8.

35. Schwartz RH, Wientzen RL, Jr., Pedreira F, Feroli EJ, Mella GW, G., et al. Penicillin V for group A streptococcal pharyngotonsillitis. A randomized trial of seven vs ten days’ therapy. JAMA. 1981;246(16):1790–5.

36. Brook I, Gober AE. Rate of eradication of group A beta-hemolytic streptococci in children with pharyngo-tonsillitis by amoxicillin and cefdinir. International Journal of Pediatric Otorhinolaryngology. 2009;73(5):757–9.

37. Schwartz RH, Kim D, Martin M, Pichichero ME. A Reappraisal of the Minimum Duration of Antibiotic Treatment Before Approval of Return to School for Children With Streptococcal Pharyngitis. Pediatric Infectious Disease Journal. 2015;34(12):1302–4.

38. Shvartzman P, Tabenkin H, Rosentzwaig A, Dolginov F. Treatment of streptococcal pharyngitis with amoxycillin once a day. BMJ. 1993;306(6886):1170–2.

39. Lester RL, Howie VM, Ploussard JH. Treatment of streptococcal pharyngitis with different antibiotic regimens. Clinical Pediatrics. 1974;13(3):239–42.

40. Trickett PC, Dineen P, Mogabgab W. Clinical experience: respiratory tract. Trimethoprim-sulfamethoxazole versus penicillin G in the treatment of group A beta-hemolytic streptococcal pharyngitis and tonsillitis. J Infect Dis. 1973;128:Suppl:693–5 p.

41. Pacifico L, Scopetti F, Ranucci A, Pataracchia M, Savignoni F, Chiesa C. Comparative efficacy and safety of 3-day azithromycin and 10-day penicillin V treatment of group A beta-hemolytic streptococcal pharyngitis in children. Antimicrob Agents Chemother. 1996;40(4):1005–8.

42. Stein GE, Christensen S, Mummaw N. Comparative study of clarithromycin and penicillin V in the treatment of streptococcal pharyngitis. Eur J Clin Microbiol Infect Dis. 1991;10(11):949–53.

43. Block SL, Hedrick JA, Tyler RD. Comparative study of the effectiveness of cefixime and penicillin V for the treatment of streptococcal pharyngitis in children and adolescents. Pediatr Infect Dis J. 1992;11(11):919–25.

44. Mogabgab WJ. Comparison of cepharadine and cephalexin in the treatment of respiratory and urinary tract infections. Curr Ther Res Clin Exp. 1976;19(4):421–32.

45. Levine MK, Berman JD. A comparison of clindamycin and erythromycin in beta--hemolytic streptococcal infections. J Med Assoc Ga. 1972;61(3):108–11.

46. Watkins VS, Smietana M, Conforti PM, Sides GD, Huck W. Comparison of dirithromycin and penicillin for treatment of streptococcal pharyngitis. Antimicrob Agents Chemother. 1997;41(1):72–5.

47. Edmond EW, Cramblett HG, Siewers CM, Crews J, Ellis B, Jenkins GR. Comparison of efficacy of phenoxymethyl penicillin and buffered penicillin G in treatment of streptococcal pharyngitis. J Pediatr. 1966;68(3):442–7.

48. Disney FA DM, Higgins JE, Nolen T, Poole JM, Randolph M, Rogan MP.. Comparison of once-daily cefadroxil and four-times-daily erythromycin in group A streptococcal tonsillopharyngitis. Advances in therapy. 1990;7(6):312–26.

49. Stillerman M. Comparison of oral cephalosporins with penicillin therapy for group A streptococcal pharyngitis. Pediatr Infect Dis. 1986;5(6):649–54.

50. Ginsburg CM, McCracken GH, Jr., Crow SD, Steinberg JB, Cope F. A controlled comparative study of penicillin V and cefadroxil therapy of group A streptococcal tonsillopharyngitis. J Int Med Res. 1980;8(Suppl 1):82–6.

51. Dagnelie CF, van der Graaf Y, De Melker RA. Do patients with sore throat benefit from penicillin? A randomized double-blind placebo-controlled clinical trial with penicillin V in general practice. Br J Gen Pract. 1996;46(411):589–93.

52. Pichichero ME, Gooch WM, Rodriguez W, Blumer JL, Aronoff SC, Jacobs RF, et al. Effective short-course treatment of acute group A beta-hemolytic streptococcal tonsillopharyngitis. Ten days of penicillin V vs 5 days or 10 days of cefpodoxime therapy in children. Arch Pediatr Adolesc Med. 1994;148(10):1053–60.

53. Ryan DC, Dreher GH, Hurst JA. Estolate and stearate forms of erythromycin in the treatment of acute beta haemolytic streptococcal pharyngitis. Med J Aust. 1973;1(1):20–1.

54. Colcher IS, Bass JW. Penicillin treatment of streptococcal pharyngitis. A comparison of schedules and the role of specific counseling. JAMA. 1972;222(6):657–9.

55. Raz R EG, Colodner R, Reiss S, Schvartzman P, Tabenkin H, and Leshem Y.. Penicillin V twice daily vs. four times daily in the treatment of streptococcal pharyngitis. Infectious Diseases in Clinical Practice. 1995;4(1):50–4.

56. Pavesio D, Pecco P, Peisino MG. Short-term treatment of streptococcal tonsillitis with ceftriaxone. Chemotherapy. 1988;34 Suppl 1:34–8.

57. Gerber MA, Randolph MF, DeMeo KK. Streptococcal antigen in the pharynx after initiation of antibiotic therapy. Pediatr Infect Dis J. 1987;6(5):489–91.

58. Sinanian R, Ruoff G, Panzer J, Atkinson W. Streptococcal pharyngitis: a comparison of the eradication of the organism by 5- and 10-day antibiotic therapy. Curr Ther Res Clin Exp. 1972;14(11):716–20.

59. Ginsburg CM, McCracken GH, Jr., Steinberg JB, Crow SD, Dildy BF, Cope F, et al. Treatment of Group A streptococcal pharyngitis in children. Results of a prospective, randomized study of four antimicrobial agents. Clin Pediatr (Phila). 1982;21(2):83–8.

60. Hoskins TW, Bernstein LS. Trimethoprim/sulphadiazine compared with penicillin V in the treatment of streptococcal throat infections. J Antimicrob Chemother. 1981;8(6):495–6.

61. Krober MS, Bass JW, Michels GN. Streptococcal pharyngitis. Placebo-controlled double-blind evaluation of clinical response to penicillin therapy. JAMA. 1985;253(9):1271–4.

62. Aronovitz GH, Morgan DL, Spitzer TQ. Streptococcal infection in pediatric patients. Comparison of dicloxacillin with phenoxymethol penicillin. Am J Dis Child. 1968;116(1):66–9.

63. Schalet N, Reen BM, Houser HB. A comparison of penicillin G and penicillin V in treatment of streptococcal sore throat. Am J Med Sci. 1958;235(2):183–8.

64. Howie VM, Ploussard JH. Treatment of group A streptococcal pharyngitis in children. Comparison of lincomycin and penicillin G given orally and benzathine penicillin G given intramuscularly. Am J Dis Child. 1971;121(6):477–80.

65. Rabinovitch M, MacKenzie R, Brazeau M, Marks MI. Treatment of streptococcal pharyngitis. I. Clinical evaluation. Can Med Assoc J. 1973;108(10):1271–4.

66. Azimi PH, Cramblett HG, Del rosario AJ, Kronfol H, Haynes RE, Hilty MD. Cephalexin: treatment of streptococcal pharyngitis. J Pediatr. 1972;80(6):1042–5.

67. De la Garza CA NT, Rogan MP. Cefprozil vs. erythromycin in tonsillopharyngitis. Infections in Medicine. 1992;9:8–20.

68. Milatovic D, Knauer J. Cefadroxil versus penicillin in the treatment of streptococcal tonsillopharyngitis. European Journal of Clinical Microbiology & Infectious Diseases. 1989;8(4):282–8.

69. Falagas ME, Vouloumanou EK, Matthaiou DK, Kapaskelis AM, Karageorgopoulos DE. Effectiveness and safety of short-course vs long-course antibiotic therapy for group a beta hemolytic streptococcal tonsillopharyngitis: a meta-analysis of randomized trials. Mayo Clin Proc. 2008;83(8):880–9.

70. Casey JR, Pichichero ME. Metaanalysis of short course antibiotic treatment for group a streptococcal tonsillopharyngitis. Pediatr Infect Dis J. 2005;24(10):909–17.

71. Altamimi S, Khalil A, Khalaiwi KA, Milner R, Pusic MV, Al Othman MA. Short versus standard duration antibiotic therapy for acute streptococcal pharyngitis in children. Cochrane Database Syst Rev. 2009(1):CD004872.

72. Holm AE, Llor C, Bjerrum L, Cordoba G. Short-vs. Long-Course Antibiotic Treatment for Acute Streptococcal Pharyngitis: Systematic Review and Meta-Analysis of Randomized Controlled Trials. Antibiotics (Basel). 2020;9(11).

73. Shulman ST, Bisno AL, Clegg HW, Gerber MA, Kaplan EL, Lee G, et al. Clinical practice guideline for the diagnosis and management of group A streptococcal pharyngitis: 2012 update by the Infectious Diseases Society of America. Clin Infect Dis. 2012;55(10):e86–102.

74. Eng RH, Smith SM, Cherubin CE, Tan EN. Evaluation of two methods for overcoming the antibiotic carry-over effect. Eur J Clin Microbiol Infect Dis. 1991;10(1):34–8.

75. Lan AJ, Colford JM, Colford JM, Jr. The impact of dosing frequency on the efficacy of 10-day penicillin or amoxicillin therapy for streptococcal tonsillopharyngitis: A meta-analysis. Pediatrics. 2000;105(2):E19.

76. Kardas P. Patient compliance with antibiotic treatment for respiratory tract infections. J Antimicrob Chemother. 2002;49(6):897–903.

77. Hoban DJ, Nauta J. Clinical And Bacteriological Impact Of Clarithromycin In Streptococcal Pharyngitis: Findings From A Meta-Analysis Of Clinical Trials. Drug Des Devel Ther. 2019;13:3551–8.

78. Gutiérrez-Castrellón P M-BJ, Bosch-Canto V, Solomon-Santibañez G, de Colsa-Ranero A.. Efficacy and safety of clarithromycin in pediatric patients with upper respiratory infections: a systematic review with meta-analysis.. Rev Invest Clin 2002;64(2):126–35.

79. Fay K, Onukwube J, Chochua S, Schaffner W, Cieslak P, Lynfield R, et al. Patterns of Antibiotic Nonsusceptibility Among Invasive Group A Streptococcus Infections-United States, 2006-2017. Clin Infect Dis. 2021;73(11):1957–64.

80. Andreoni F, Zurcher C, Tarnutzer A, Schilcher K, Neff A, Keller N, et al. Clindamycin Affects Group A Streptococcus Virulence Factors and Improves Clinical Outcome. J Infect Dis. 2017;215(2):269–77.

81. Stevens DL, Bisno AL, Chambers HF, Everett ED, Dellinger P, Goldstein EJ, et al. Practice guidelines for the diagnosis and management of skin and soft-tissue infections. Clin Infect Dis. 2005;41(10):1373–406.

82. Excellence NIfC. Drug allergy: diagnosis and management 2014.

83. Prevention CfDCa. Is it really penicillin allergy? 2017.

84. Imohl M, van der Linden M. Antimicrobial Susceptibility of Invasive Streptococcus pyogenes Isolates in Germany during 2003-2013. PLoS One. 2015;10(9):e0137313.

85. Pichichero ME, Casey JR. Systematic review of factors contributing to penicillin treatment failure in Streptococcus pyogenes pharyngitis. Otolaryngol Head Neck Surg. 2007;137(6):851–7.

86. Brook I. The role of beta-lactamase producing bacteria and bacterial interference in streptococcal tonsillitis. Int J Antimicrob Agents. 2001;17(6):439–42.

87. Schaad UB. Acute streptococcal tonsillopharyngitis: a review of clinical efficacy and bacteriological eradication. J Int Med Res. 2004;32(1):1–13.

