## Supplementary Figure S1 for "Time to negative throat culture following initiation of antibiotics for pharyngeal group A *Streptococcus*: a systematic review and meta-analysis to inform public health control measures"

### Figure S1. Penicillin V - proportion of patients with culture-confirmed group A streptococcal (GAS) throat infection or carriage at day 1 (24 hours after the start of antibiotics), day 2 and days 3-9 (n=28 studies).

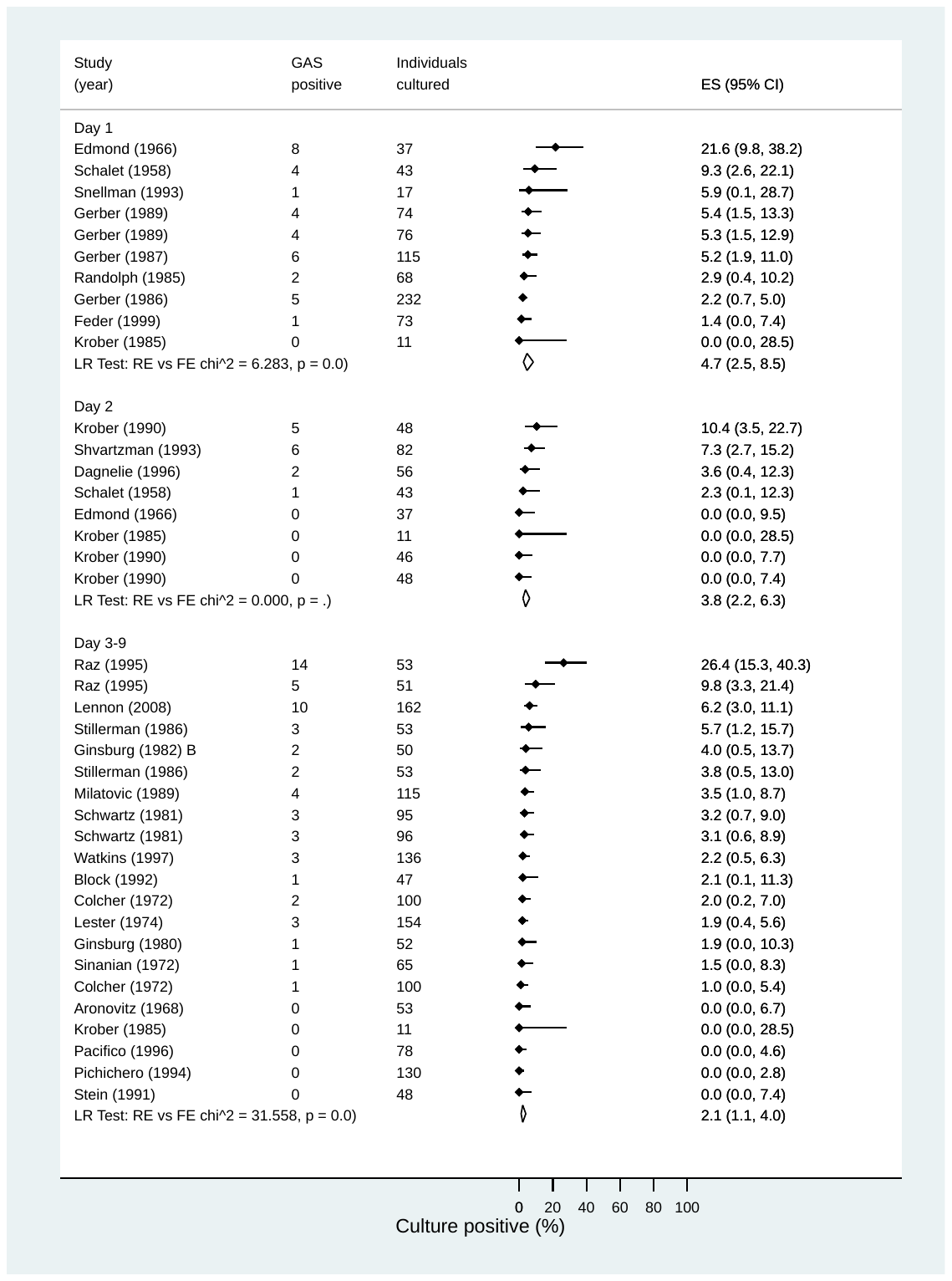
