## Supplementary Figure S2 for "Time to negative throat culture following initiation of antibiotics for pharyngeal group A *Streptococcus*: a systematic review and meta-analysis to inform public health control measures"

### Figure S2. All antibiotics - meta-regression of proportion of patients with culture-confirmed group A streptococcal (GAS) throat infection or carriage while on antibiotics (days 1 to 9 after the start of antibiotics) (n=42 studies).

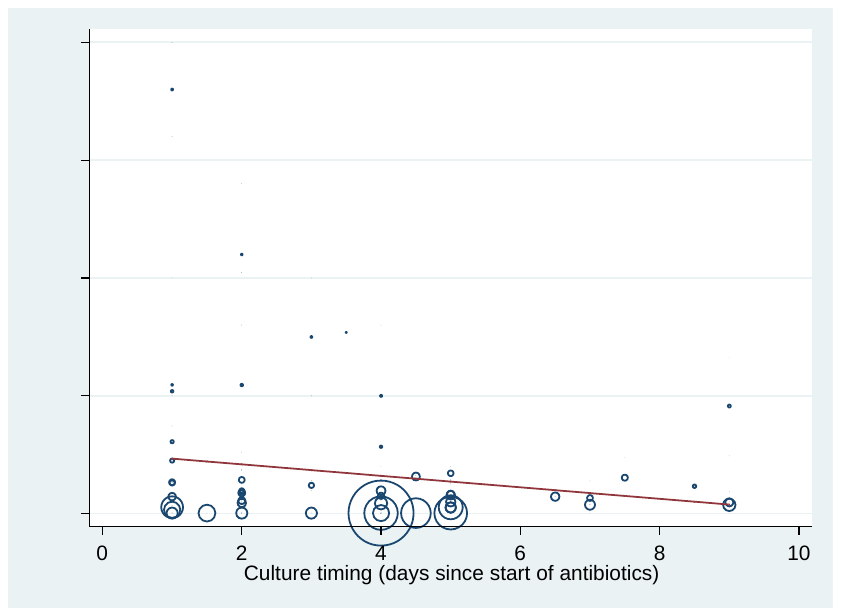
