## Supplementary Figure S6 for "Time to negative throat culture following initiation of antibiotics for pharyngeal group A *Streptococcus*: a systematic review and meta-analysis to inform public health control measures"

### Figure S6. Proportion of patients with culture-confirmed group A streptococcal (GAS) throat infection or carriage at different times after completing antibiotic treatment (Early <72hrs, Intermediate 72hrs-10 days, Late >10 days) (n=23 studies).

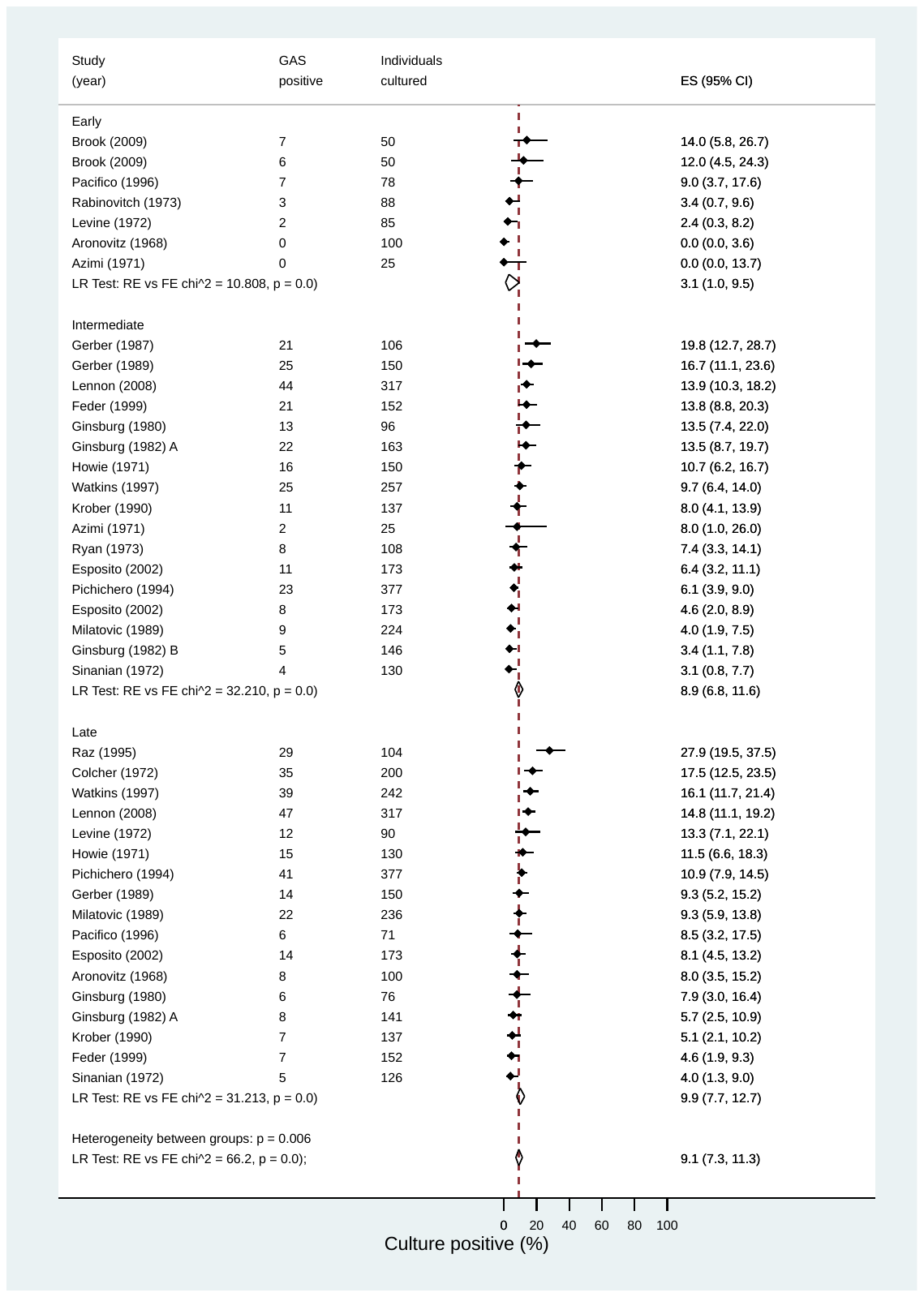
