## Supplementary Figure S7 for "Time to negative throat culture following initiation of antibiotics for pharyngeal group A *Streptococcus*: a systematic review and meta-analysis to inform public health control measures"

### Figure S7. Proportion of patients with relapse or reacquisition of the original group A Streptococcal (GAS) strain after completion of therapy by antibiotic class, where typing was reported (n=13 studies).

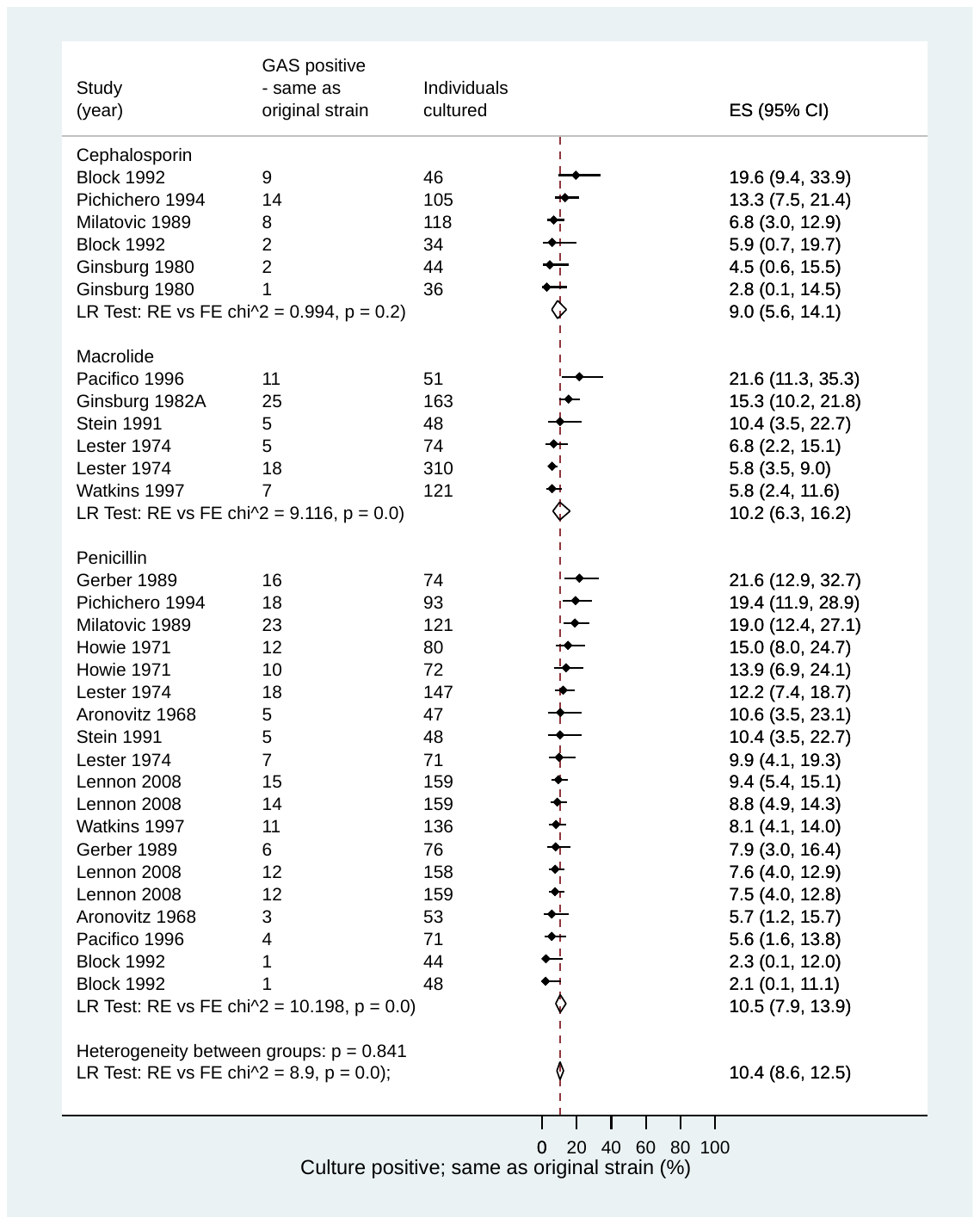
