## Supplementary Figure S8 for "Time to negative throat culture following initiation of antibiotics for pharyngeal group A *Streptococcus*: a systematic review and meta-analysis to inform public health control measures"

### Figure S8. Proportion of patients with acquisition of a new strain of group A Streptococcal (GAS) after completion of therapy by antibiotic class, where typing was reported (n=13 studies).

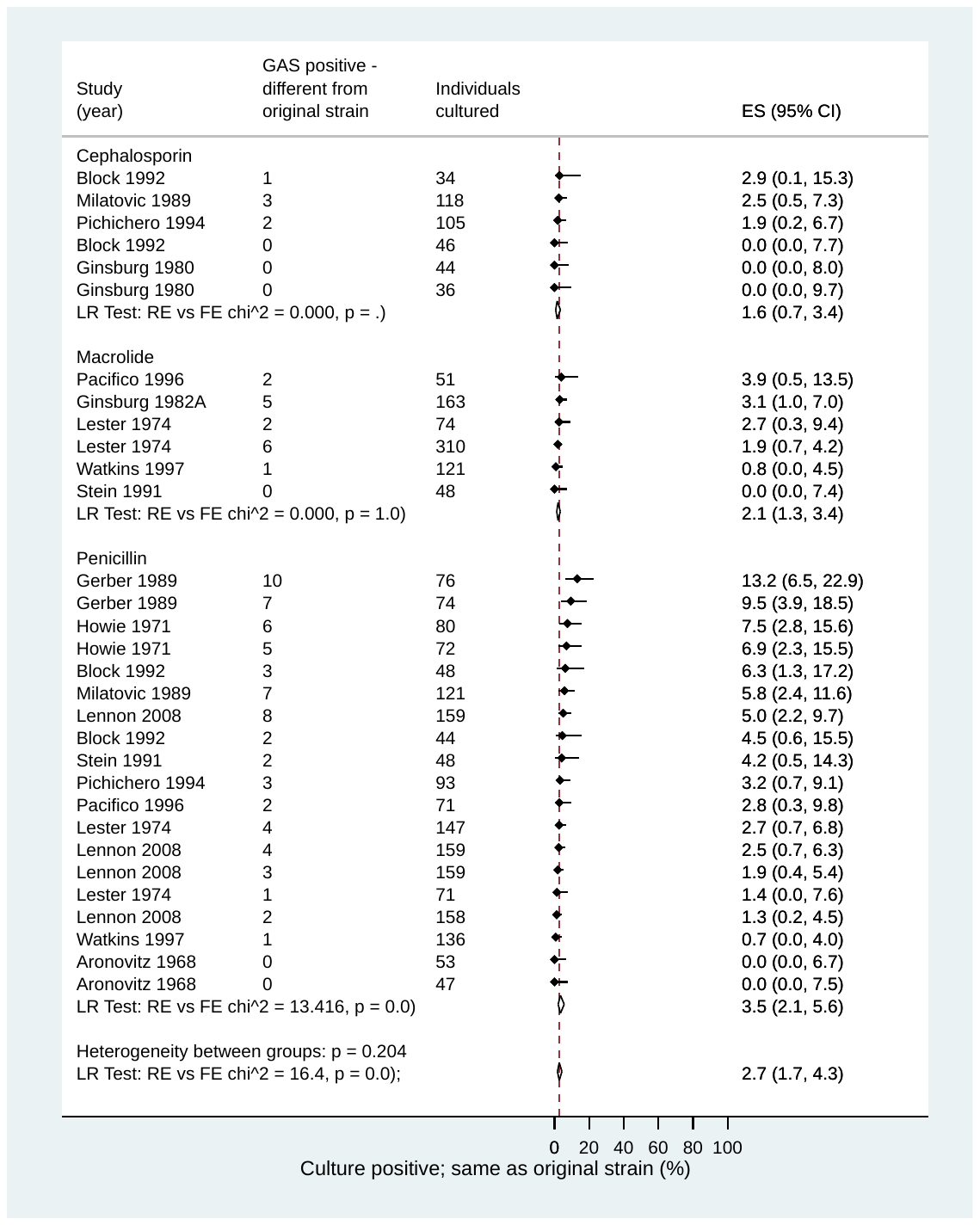
