## Supplementary Figure S9 for "Time to negative throat culture following initiation of antibiotics for pharyngeal group A *Streptococcus*: a systematic review and meta-analysis to inform public health control measures"

### Figure S9. Proportion of patients who reported any side effect or adverse drug reaction by antibiotic class, where reported (n=14 studies).

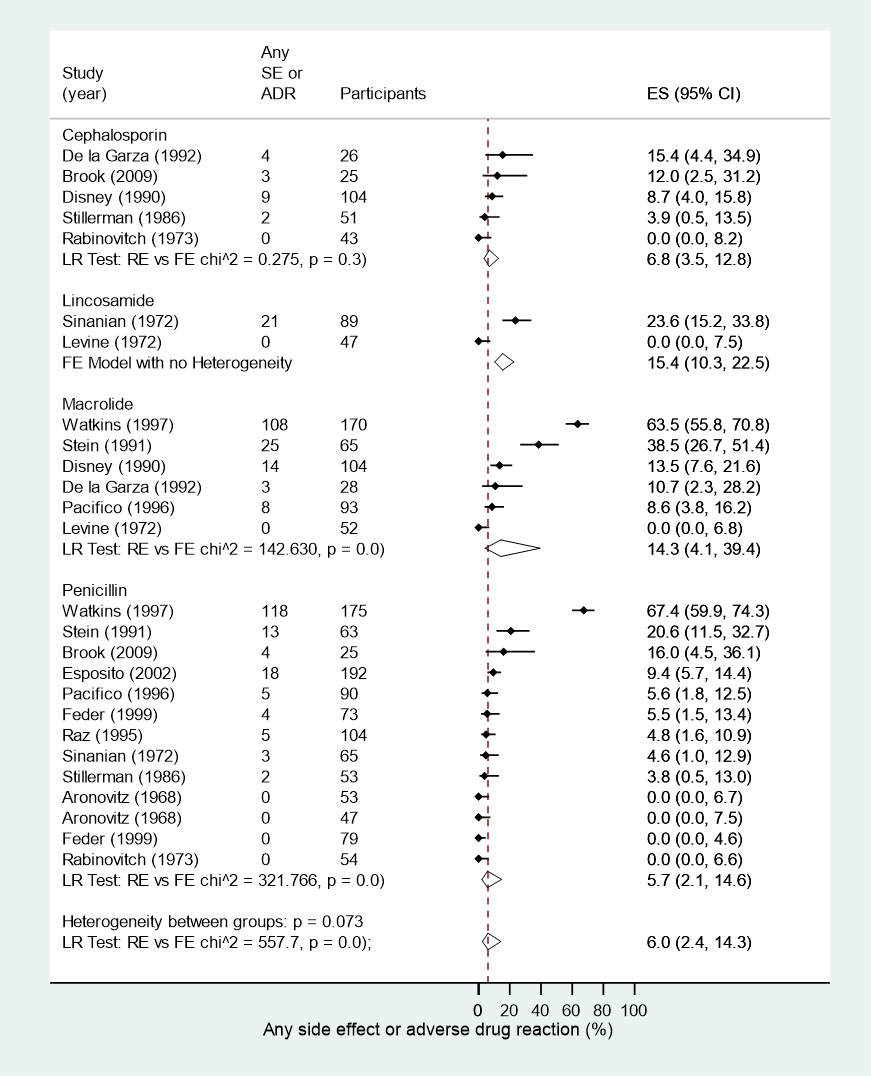
