## Supplementary Figure S10 for "Time to negative throat culture following initiation of antibiotics for pharyngeal group A *Streptococcus*: a systematic review and meta-analysis to inform public health control measures"

### Figure S10. Proportion of patients who ceased the study drug due to a side effect or adverse drug reaction by antibiotic class, where reported (n=12 studies).

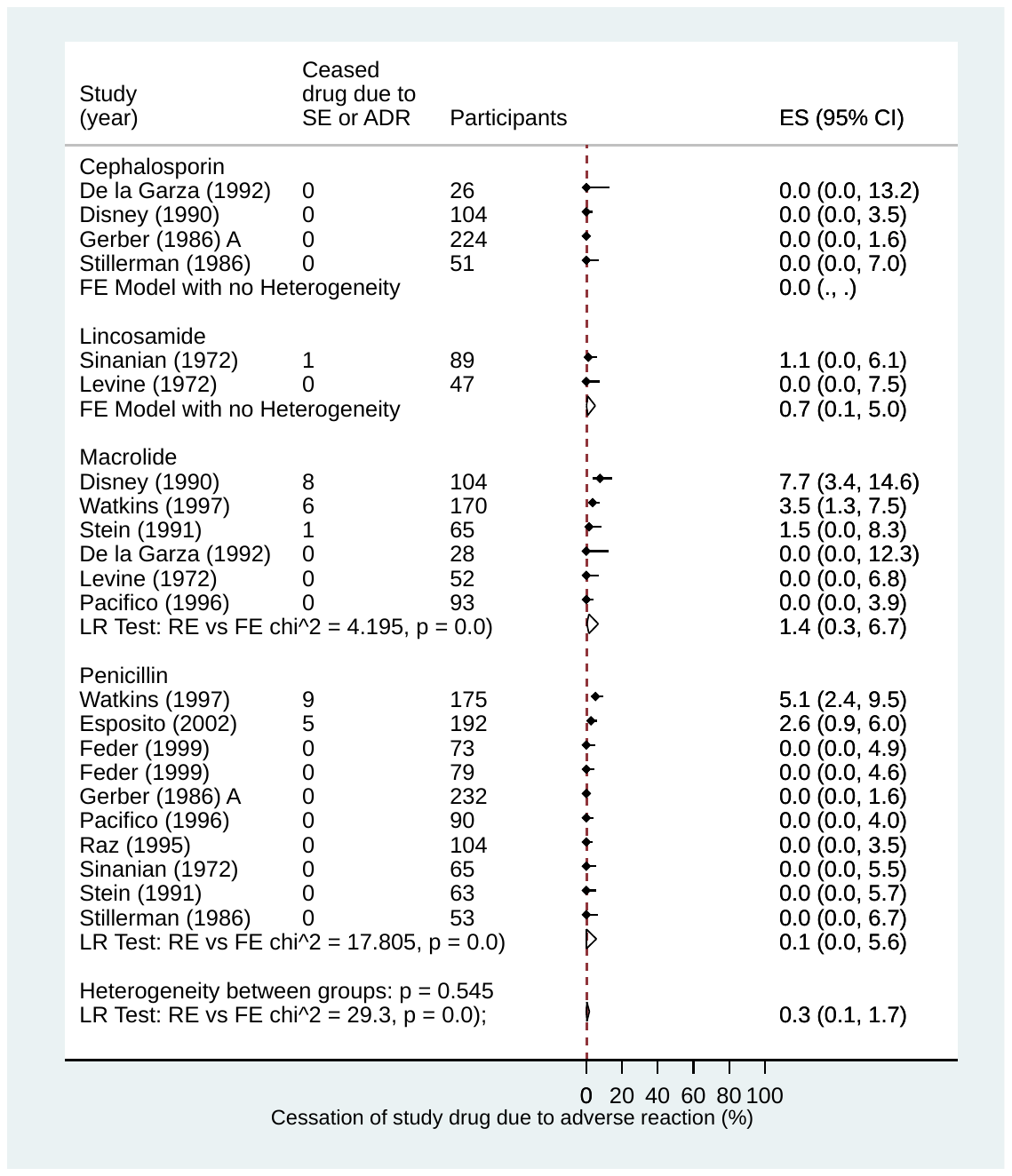
