## Supplementary Appendix A for "Time to negative throat culture following initiation of antibiotics for pharyngeal group A *Streptococcus*: a systematic review and meta-analysis to inform public health control measures"

### Supplementary Appendix A. Search strategy

Database: Ovid MEDLINE(R) ALL <1946 to October 18, 2021>

Search Strategy:

--------------------------------------------------------------------------------

1	streptococcal infections/ or rheumatic fever/ or scarlet fever/ (45136)

2	"streptococc* infection* ".tw,kw. (6645)

3	"scarlet fever".kw,tw. (2367)

4	pharyngitis.kw,tw. (6051)

5	pyogenes.kw,tw. (10230)

6	(strep* adj2 Carriage).kw,tw. (526)

7	or/1-6 (58878)

8	exp Anti-Bacterial Agents/ (764976)

9	exp Penicillins/ (82399)

10	Azithromycin/ (6120)

11	Cefaclor/ (841)

12	Cefadroxil/ (417)

13	cefdinir/ (252)

14	Cefixime/ (794)

15 Cefotiam/ (414)

16	Ceftizoxime/ (1148)

17	Ceftibuten/ (215)

18	Cefuroxime/ (2246)

19	exp Cephalosporins/ (44226)

20	Clarithromycin/ (6572)

21	clindamycin/ (5882)

22	erythromycin/ (13955)

23	josamycin/ (235)

24	lincosamide/ (465)

25	loracarbef/ (0)

26	macrolide/ (13124)

27	penicillin derivative/ (0)

28	penicillin V/ (2180)

29	roxithromycin/ (836)

30	spiramycin/ (725)

31	telithromycin/ (0)

32	Amoxicillin/ (9913)

33	Amoxicillin-Potassium Clavulanate Combination/ (2674)

34	Clavulanic Acid/ (1678)

35	Quinolones/ (12329)

36	Moxifloxacin/ (2692)

37	Gemifloxacin/ (278)

38	Levofloxacin/ (3629)

39	(azithromycin or cefaclor or cefadroxil or cefcapene or cefdinir or cefixime or cefotiam or cefpodoxime or ceftibuten or cefuroxime or cephalosporin or clarithromycin or "clavulanic acid" or clindamycin or erythromycin or josamycin or lincosamide or loracarbef or macrolide or penicillin or roxithromycin or spiramycin or telithromycin).kw,tw. (119821)

40	(Amoxicillin or Augmentin or Co-amoxiclav or Clavulanate or Quinolone or Moxifloxacin or Gemifloxacin or Levofloxacin).kw,tw. (38019)

41	exp Antibiotic Prophylaxis/ (14723)

42	Chemoprevention/ (6331)

43	(Prophylaxis or Chemoprophylaxis).kw,tw. (107937)

44	or/8-43 (911843)

45	clinical effectiveness/ (1060749)

46	intervention study/ (531530)

47	time factor/ (1217374)

48	outcome assessment/ (0)

49	Outcome Assessment, Health Care/ (77921)

50	treatment outcome/ (1060749)

51	"eradicat*".kw,tw. (68818)

52	Clearance.kw,tw. (172556)

53	or/45-52 (2815855)

54	7 and 44 and 53 (2779)

55	limit 54 to english language (2216)

56	limit 55 to humans (1998)

57	(letter or historical article or comment or editorial or news).pt. (2537403)

58	56 not 57 (1910)

***************************

Database: Embase <1974 to 2021 October 25>

Search Strategy:

--------------------------------------------------------------------------------

1	exp streptococcus infection/ or exp group a streptococcal infection/ (51972)

2	exp rheumatic fever/ (7775)

3	exp scarlet fever/ (1453)

4	"streptococc* infection*".kw,tw. (5851)

5	"scarlet fever".kw,tw. (1078)

6	pharyngitis.kw,tw. (7924)

7	pyogenes.kw,tw. (11636)

8	(strep* adj2 Carriage).kw,tw. (624)

9	or/1-8 (69197)

10	exp antiinfective agent/ (3798679)

11	exp penicillin derivative/ (313240)

12	azithromycin/ (43793)

13	cefaclor/ (8195)

14	cefadroxil/ (3657)

15	cefdinir/ (2386)

16	cefixime/ (8447)

17	cefotiam/ (3186)

18	ceftizoxime/ (4107)

19	ceftibuten/ (1381)

20	cefuroxime axetil/ or cefuroxime/ (27696)

21	exp cephalosporin derivative/ (247374)

22	amoxicillin plus clarithromycin plus omeprazole/ or clarithromycin/ or exp clarithromycin derivative/ or amoxicillin plus clarithromycin plus lansoprazole/ (39249)

23	clindamycin/ (54805)

24	erythromycin/ or exp erythromycin derivative/ (75866)

25	josamycin/ (2308)

26	lincosamide/ (2997)

27	loracarbef/ (1072)

28	macrolide/ (33829)

29	roxithromycin/ (5738)

30	spiramycin/ (4448)

31	telithromycin/ (2942)

32	amoxicillin plus clavulanic acid/ or amoxicillin/ or exp amoxicillin derivative/ (98808)

33	clavulanic acid/ (14642)

34	exp quinolone derivative/ (186769)

35	moxifloxacin/ (20882)

36	gemifloxacin/ (1589)

37	levofloxacin/ (41168)

38	(azithromycin or cefaclor or cefadroxil or cefcapene or cefdinir or cefixime or cefotiam or cefpodoxime or ceftibuten or cefuroxime or cephalosporin or clarithromycin or "clavulanic acid" or clindamycin or erythromycin or josamycin or lincosamide or loracarbef or macrolide or penicillin or roxithromycin or spiramycin or telithromycin).kw,tw. (139399)

39	(Amoxicillin or Augmentin or Co-amoxiclav or Clavulanate or Quinolone or Moxifloxacin or Gemifloxacin or Levofloxacin).kw,tw. (59308)

40	Antibiotic Prophylaxis.kw,tw. (15251)

41	exp antibiotic prophylaxis/ (34332)

42	exp chemoprophylaxis/ (26282)

43	(Prophylaxis or Chemoprophylaxis).kw,tw. (163016)

44	or/10-43 (3947417)

45	exp comparative effectiveness/ or exp clinical effectiveness/ (255315)

46	exp intervention study/ (52425)

47	exp time factor/ (42129)

48	exp outcome assessment/ (629063)

49	exp treatment outcome/ (1864693)

50	outcome.ti. (240561)

51	"eradicat*".kw,tw. (91853)

52	Clearance.kw,tw. (235348)

53	or/45-52 (2529184)

54	9 and 44 and 53 (4030)

55	(54 and english.lg.) not (letter or editorial).pt. not (nonhuman/ not human/) not (conference abstract or conference paper or conference proceeding or "conference review").pt. (2808)

***************************

Search Name:	20211101MCPenicillinTreatmentStreptococcalInfections58989

Date Run:	26/10/2021 14:51:04

Comment:

ID	Search	Hits

#1	MeSH descriptor: [Streptococcal Infections] explode all trees	1533

#2	MeSH descriptor: [Rheumatic Fever] explode all trees	188

#3	MeSH descriptor: [Scarlet Fever] explode all trees	12

#4	(streptococc* AND infection*):ti,ab,kw	2919

#5	("scarlet fever"):ti,ab,kw	55

#6	(pharyngitis):ti,ab,kw	2468

#7	(pyogenes):ti,ab,kw	502

#8	(strep* AND Carriage):ti,ab,kw	243

#9	#1 OR #2 OR #3 OR #4 OR #5 OR #6 OR #7 OR #8	5729

#10	MeSH descriptor: [Anti-Bacterial Agents] explode all trees	12590

#11	MeSH descriptor: [Penicillins] explode all trees	5783

#12	MeSH descriptor: [Azithromycin] explode all trees	1052

#13	MeSH descriptor: [Cefaclor] explode all trees	230

#14	MeSH descriptor: [Cefadroxil] explode all trees	94

#15	MeSH descriptor: [Cefdinir] explode all trees	64

#16	MeSH descriptor: [Cefixime] explode all trees	145

#17	MeSH descriptor: [Cefotiam] explode all trees	46

#18	MeSH descriptor: [Ceftizoxime] explode all trees	174

#19	MeSH descriptor: [Ceftibuten] explode all trees	42

#20	MeSH descriptor: [Cefuroxime] explode all trees	468

#21	MeSH descriptor: [Cephalosporins] explode all trees	4439

#22	MeSH descriptor: [Clarithromycin] explode all trees	1510

#23	MeSH descriptor: [Clindamycin] explode all trees	893

#24	MeSH descriptor: [Erythromycin] explode all trees	3485

#25	MeSH descriptor: [Josamycin] explode all trees	21

#26	MeSH descriptor: [Lincosamides] explode all trees	935

#27	MeSH descriptor: [Macrolides] explode all trees	9459

#28	MeSH descriptor: [Roxithromycin] explode all trees	120

#29	MeSH descriptor: [Spiramycin] explode all trees	29

#30	MeSH descriptor: [Amoxicillin] explode all trees	2914

#31	MeSH descriptor: [Clavulanic Acids] explode all trees	900

#32	MeSH descriptor: [Quinolones] explode all trees	5162

#33	MeSH descriptor: [Moxifloxacin] explode all trees	882

#34	MeSH descriptor: [Gemifloxacin] explode all trees	47

#35	MeSH descriptor: [Levofloxacin] explode all trees	655

#36	(azithromycin or cefaclor or cefadroxil or cefcapene or cefdinir or cefixime or cefotiam or cefpodoxime or ceftibuten or cefuroxime or cephalosporin or clarithromycin or clavulanic acid or clindamycin or erythromycin or josamycin or lincosamide or loracarbef or macrolide or penicillin or roxithromycin or spiramycin or telithromycin):ti,ab,kw	15873

#37	(Amoxicillin or Augmentin or Co-amoxiclav or Clavulanate or Quinolone or Moxifloxacin or Gemifloxacin or Levofloxacin):ti,ab,kw	9426

#38	MeSH descriptor: [Antibiotic Prophylaxis] explode all trees	1315

#39	(Prophylaxis or Chemoprophylaxis):ti,ab,kw	27481

#40	#10 or #11 or #12 or #13 or #14 or #15 or #16 or #17 or #18 or #19 or #20 or #21 or #22 or #23 or #24 or #25 or #26 or #27 or #28 or #29 or #30 or #31 or #32 or #33 or #34 or #35 or #36 or #37 or #38 or #39	63223

#41	#9 AND #40	2107

#42	MeSH descriptor: [Treatment Outcome] explode all trees	146733

#43	(outcome or intervention or effective* or eridcat* or clearance):ti,ab,kw	870925

#44	#42 or #43	874825

#45	#41 and #44	1200
