## Supplementary Appendix B for "Time to negative throat culture following initiation of antibiotics for pharyngeal group A *Streptococcus*: a systematic review and meta-analysis to inform public health control measures"

### Supplementary Appendix B. Inclusion and exclusion criteria

|  | **Inclusion criteria** | **Exclusion criteria** |
| --- | --- | --- |
| **Population** | - Culture confirmed GAS pharyngitis or scarlet fever or with asymptomatic pharyngeal GAS carriage† | - Studies which did not report on participants with pharyngeal GAS (including those with impetigo, erysipelas, or iGAS e.g. necrotising fasciitis, pneumonia, and bacteraemia) unless a sub-group with culture confirmed pharyngeal GAS were also reported. - Studies which only reported treatment outcomes in patients with recurrent GAS pharyngitis (as we considered this to be a sub-population which may have issues affecting treatment efficacy such as penicillin tolerance, compliance issues or beta-lactamase producing co-pathogens). |
| **Intervention** | - Any antibiotic | - Studies which only reported on the use of herbal medicines or probiotics. |
| **Outcome** | - Rates of positive or negative GAS throat culture at defined time points during antibiotics or time to clearance (mean or median) after the initiation of antibiotics | - Studies which only reported bacteriological outcomes after the completion of antibiotic treatment - Studies that included the study population as a sub-group, but which do not present disaggregated data for the outcome measure. |
| **Study type** | - All peer reviewed primary research studies with 10 or more participants were considered regardless of design. | - Studies which only reported PCR or RADT (without throat culture) or which did not differentiate GAS from beta-haemolytic streptococci. - Animal or in vitro studies. - Case reports, letters, commentaries, and conference abstracts. - Systematic reviews and meta-analyses were excluded but any reviews identified during the title and abstract screening were used to screen for further eligible studies. - Duplicate publications of the same raw data. |

GAS: group A *Streptococcus*; iGAS: invasive GAS infection; PCR: polymerase chain reaction; RADT: rapid antigen detection test

† Acceptable confirmation methods were those listed in the WHO publication “Laboratory diagnosis of group A streptococcal infections”^[1]^ Studies which did not state method of laboratory confirmation but reported diagnosis of group A streptococcal infection (rather than unqualified ‘Streptococcal infection’ alone) were also included.

1. Johnson DR KE, Sramek J, Bicova R, Havlicek J, Havlickova H, Motlova J, Kriz P; for World Health Organisation Geneva. Laboratory diagnosis of group A streptococcal infections. 1996.
