## Supplementary Appendix C for "Time to negative throat culture following initiation of antibiotics for pharyngeal group A *Streptococcus*: a systematic review and meta-analysis to inform public health control measures"

### Supplementary Appendix C. Risk of bias assessments of included controlled intervention studies

| **NIHR Tool - Quality Assessment of Controlled Intervention Studies** | | | | | | | | | | | | | | |
| --- | --- | --- | --- | --- | --- | --- | --- | --- | --- | --- | --- | --- | --- | --- |
| **Author year** | **Q1** | **Q2** | **Q3** | **Q4** | **Q5** | **Q6** | **Q7** | **Q8** | **Q9** | **Q10** | **Q11** | **Q12** | **Q13** | **Q14** |
| Esposito 2002 | Y | Y | Y | N | Y | Y | Y | Y | Y | Y | Y | NR | Y | Y |
| Gerber 1986A | Y | CD | NR | NR | NR | NR | Y | Y | Y | NR | Y | NR | Y | CD |
| Snellman 1993 | Y | Y | N | N | N | NR | Y | CD | Y | Y | Y | NR | N | NR |
| Randolph 1985 | Y | Y | Y | Y | Y | Y | Y | Y | Y | Y | Y | NR | Y | CD |
| Gerber 1989 | Y | CD | CD | NR | NR | N | Y | CD | N | NR | Y | NR | Y | N |
| Ginsburg 1982A | Y | CD | NR | NR | Y | Y | Y | Y | Y | NR | Y | NR | Y | N |
| Lennon 2008 | Y | Y | Y | N | NR | Y | Y | Y | Y | NR | Y | Y | Y | N |
| Feder 1999 | Y | Y | Y | N | Y | Y | Y | Y | N | NR | Y | NR | Y | N |
| Krober 1990 | Y | Y | NR | NR | NR | Y | Y | CD | CD | NR | Y | NR | Y | N |
| Schwartz 1981 | Y | CD | NR | N | NR | Y | Y | CD | CD | NR | Y | NR | Y | N |
| Schwartz 2015 | Y | Y | NR | N | N | Y | Y | CD | Y | NR | Y | NR | Y | N |
| Shvartzman 1993 | Y | CD | NR | N | NR | Y | CD | CD | NR | NR | CD | NR | Y | N |
| Lester 1974 | Y | Y | Y | N | NR | Y | Y | Y | CD | NR | Y | NR | Y | CD |
| Trickett 1973 | Y | CD | Y | Y | Y | Y | Y | CD | NR | Y | CD | NR | Y | N |
| Pacifico 1996 | Y | Y | NR | N | NR | Y | Y | Y | Y | CD | Y | NR | Y | N |
| Stein 1991 | Y | Y | NR | Y | NR | Y | Y | CD | NR | Y | Y | NR | Y | N |
| Block 1992 | Y | Y | NR | N | N | Y | Y | Y | Y | Y | Y | NR | Y | N |
| Mogabgab 1976 | N | N | NR | NR | NR | N | NR | NR | NR | NR | Y | NR | Y | Y |
| Levine 1972 | Y | CD | NR | Y | Y | Y | Y | Y | NR | NR | Y | NR | Y | N |
| Watkins 1997 | Y | Y | Y | Y | Y | Y | Y | Y | NR | NR | Y | NR | Y | N |
| Edmond 1966 | N | NR | NR | Y | Y | NR | CD | CD | NR | NR | Y | NR | Y | NR |
| Disney 1990 | Y | CD | NR | N | N | Y | Y | CD | Y | Y | Y | NR | Y | N |
| Stillerman 1986 | Y | Y | N | N | N | Y | Y | CD | Y | NR | Y | NR | Y | N |
| Ginsburg 1980 | Y | CD | NR | N | N | Y | Y | Y | CD | NR | Y | NR | Y | N |
| Dagnelie 1996 | Y | CD | NR | Y | Y | CD | Y | Y | NR | Y | Y | NR | Y | Y |
| Pichichero 1994 | Y | CD | NR | N | Y | Y | Y | CD | Y | Y | Y | NR | Y | N |
| Ryan 1973 | Y | CD | Y | NR | Y | NR | CD | CD | NR | Y | Y | NR | Y | CD |
| Colcher 1972 | Y | CD | NR | N | N | Y | Y | CD | N | NR | Y | NR | Y | N |
| Raz 1995 | Y | CD | NR | N | N | Y | Y | Y | N | NR | Y | NR | Y | N |
| Pavesio 1988 | Y | CD | NR | N | N | Y | CD | CD | NR | NR | CD | NR | Y | CD |
| Sinanian 1972 | Y | NR | CD | N | N | CD | Y | Y | NR | NR | CD | NR | N | Y |
| Ginsburg 1982B | Y | CD | NR | N | N | Y | NR | NR | Y | NR | Y | NR | Y | NR |
| Hoskins 1981 | Y | CD | NR | N | N | NR | CD | CD | NR | NR | N | NR | Y | N |
| Krober 1985 | Y | Y | Y | Y | Y | Y | Y | Y | Y | Y | Y | NR | Y | Y |
| Aronovitz 1968 | Y | Y | Y | Y | Y | CD | Y | Y | N | Y | Y | NR | Y | Y |
| Schalet 1958 | N | NR | NR | N | Y | CD | Y | Y | Y | Y | Y | NR | CD | NR |
| Howie 1971 | Y | Y | NR | N | N | CD | CD | CD | Y | Y | Y | NR | Y | CD |
| Rabinovitch 1973 | Y | CD | NR | NR | NR | NR | N | CD | Y | NR | Y | NR | Y | N |
| Milatovic 1989 | Y | CD | N | NR | NR | CD | Y | NR | CD | NR | Y | NR | Y | N |
| De La Garza 1992 | Y | Y | NR | N | N | N | N | Y | Y | Y | Y | NR | Y | N |

Y: yes; N: no; CD: cannot determine; NR: not reported

Q1-14: questions 1 to 14 for each NIHR tool:^[1]^

Q1: Was the study described as randomized, a randomized trial, a randomized clinical trial, or an RCT?

Q2: Was the method of randomization adequate (i.e., use of randomly generated assignment)?

Q3: Was the treatment allocation concealed (so that assignments could not be predicted)?

Q4: Were study participants and providers blinded to treatment group assignment?

Q5: Were the people assessing the outcomes blinded to the participants' group assignments?

Q6: Were the groups similar at baseline on important characteristics that could affect outcomes (e.g., demographics, risk factors, co-morbid conditions)?

Q7: Was the overall drop-out rate from the study at endpoint 20% or lower of the number allocated to treatment?

Q8: Was the differential drop-out rate (between treatment groups) at endpoint 15 percentage points or lower?

Q9: Was there high adherence to the intervention protocols for each treatment group?

Q10: Were other interventions avoided or similar in the groups (e.g., similar background treatments)?

Q11: Were outcomes assessed using valid and reliable measures, implemented consistently across all study participants?

Q12: Did the authors report that the sample size was sufficiently large to be able to detect a difference in the main outcome between groups with at least 80% power?

Q13: Were outcomes reported or subgroups analysed prespecified (i.e., identified before analyses were conducted)?

Q14: Were all randomized participants analysed in the group to which they were originally assigned, i.e., did they use an intention-to-treat analysis?

| **NIHR Tool – Quality Assessment of for Before-After (Pre-Post) Studies with No Control Group** | | | | | | | | | | | | |
| --- | --- | --- | --- | --- | --- | --- | --- | --- | --- | --- | --- | --- |
| **Author year** | **Q1** | **Q2** | **Q3** | **Q4** | **Q5** | **Q6** | **Q7** | **Q8** | **Q9** | **Q10** | **Q11** | **Q12** |
| Brook 2009 | Y | Y | Y | CD | NR | Y | Y | N | Y | N | N | NR |
| Azimi 1971 | Y | N | NR | CD | NR | Y | Y | N | N | N | N | NR |
| Gerber 1987 | Y | N | CD | CD | NR | N | Y | N | N | N | N | NR |

Y: yes; N: no; CD: cannot determine; NR: not reported

Q1-12: questions 1 to 14 for each NIHR tool:^1^

Q1: Was the study question or objective clearly stated?

Q2: Were eligibility/selection criteria for the study population prespecified and clearly described?

Q3: Were the participants in the study representative of those who would be eligible for the test/service/intervention in the general or clinical population of interest?

Q4: Were all eligible participants that met the prespecified entry criteria enrolled?

Q5: Was the sample size sufficiently large to provide confidence in the findings?

Q6: Was the test/service/intervention clearly described and delivered consistently across the study population?

Q7: Were the outcome measures prespecified, clearly defined, valid, reliable, and assessed consistently across all study participants?

Q8: Were the people assessing the outcomes blinded to the participants' exposures/interventions?

Q9: Was the loss to follow-up after baseline 20% or less? Were those lost to follow-up accounted for in the analysis?

Q10: Did the statistical methods examine changes in outcome measures from before to after the intervention? Were statistical tests done that provided p values for the pre-to-post changes?

Q11: Were outcome measures of interest taken multiple times before the intervention and multiple times after the intervention (i.e., did they use an interrupted time-series design)?

Q12: If the intervention was conducted at a group level (e.g., a whole hospital, a community, etc.) did the statistical analysis take into account the use of individual-level data to determine effects at the group level?

1. Health NIo. Study quality assessment tools. . Bethesda: National Institutes of Health; 2020
