## Supplementary Appendix D for "Time to negative throat culture following initiation of antibiotics for pharyngeal group A *Streptococcus*: a systematic review and meta-analysis to inform public health control measures"

### Supplementary Appendix D. Proportion of patients with culture-confirmed group A streptococcal (GAS) throat carriage in studies reporting on macrolides, lincosamides and sulphonamides (n=15).

| **Study (Reference)** | **Antibiotic regimen** | **Day of culture** | **Proportion culture-positive** |
| --- | --- | --- | --- |
| **Macrolides** | | | |
| Snellman 1993^[1]^ | Erythromycin 250mg TDS | Day 1 | 6/15 (40.0%) |
| Ryan 1973^[2]^ | Erythromycin 30-50mg/kg/day QDS | Day 1-3 | 2/110 (1.8%) |
| Levine 1972^[3]^ | Erythromycin 16mg/kg/day TDS/QDS | Day 2-6 | 3/52 (5.8%) |
| Watkins 1997^[4]^ | Dirithromycin 500mg OD | Day 3-5 | 7/121 (5.8%) |
| Stein 1991^[5]^ | Clarithromycin 250mg BD | Day 4-6 | 0/47 (0%) |
| Ginsburg 1982A^[6]^ | Erythromycin 15mg/kg/day BD | Day 5 | 12/175 (6.9%) |
| Lester 1974^[7]^ | Erythromycin 500mg/day if <22.6kg, 1g/day if >22.7kg QDS | Day 5 | 2/74 (2.7%) |
| Ginsburg 1982B^[8]^ | Erythromycin 15mg/kg/day BD | Day 5 | 0/50 (0%) |
| Disney 1990^[9]^ | Erythromycin 30mg/kg/day QDS | Day 7-8 | 8/84 (9.5%) |
| De la Garza 1992^[10]^ | Erythromycin 30mg/kg/day OD | Day 7-10 | 1/22 (4.5%) |
| **Lincosamides** | | | |
| Levine 1972^3^ | Clindamycin 16mg/kg/day TDS/QDS | Day 2-6 | 0/47 (0%) |
| Lester 1974^7^ | Clindamycin palmitate <24.9kg 300mg/day QDS, 25-34kg 450mg/day TDS, >34.1 600mg/day QDS, Clindamycin HCL <24.9kg 300mg/day QDS, 25-34kg 450mg/day TDS, >34.1 600mg/day QDS or Clindamycin HCL <24.9kg 300mg/day >25kg 600mg/day BD | Day 5 | 1/323 (0.3%) |
| Sinanian 1972^[11]^ | Clindamycin Up to 55lb 75mg QDS to 150mg TDS, 55lb-75lb 150mg TDS to 150mg QDS, >75lb 150mg QDS to 300mg QDS | Day 7 | 1/67 (1.5%) |
| **Sulphonamides** | | | |
| Trickett 1973^[12]^ | Co-trimoxazole 2 tablets BD (80/400) | Day 2 | 18/44 (40.9%) |
| Hoskins 1981^[13]^ | Co-trimoxazole 2 tablets BD (125/375) | Day 3-4 | 4/13 (30.8%) |
