## Supplementary Appendix E for "Time to negative throat culture following initiation of antibiotics for pharyngeal group A *Streptococcus*: a systematic review and meta-analysis to inform public health control measures"

### Supplementary Appendix E. Evidence of heterogeneity and of differences between sub-groups

|  | **LR test statistic** | **Degrees of Freedom** | **p-value** | **Tau^2^** |
| --- | --- | --- | --- | --- |
| **Figure 2**  Day 1  Day 2  Day 3-9  Test for heterogeneity between sub-groups | 180.0  58.3  120.2  2.7 | 9  7  29  2 | <0.01  <0.01  <0.01  0.3 | 2.6  1.7  1.4  - |
| **Figure 3**  Day 1  Day 2  Day 3-9  Test for heterogeneity between sub-groups | 95.8  33.8  74.9  2.0 | 13  11  26  2 | <0.01  <0.01  <0.01  0.4 | 2.5  1.7  1.4  - |
| **Figure 4**  Cephalosporin  Macrolide  Penicillin  Overall  Test for heterogeneity between sub-groups | 1.1  10.2  26.0  29.7  1.6 | 4  4  17  29  2 | 0.1  <0.01  <0.01  <0.01  0.5 | 0.1  0.3  0.2  0.2  - |
| **Figure S1**  Day 1  Day 2  Day 3-9  Test for heterogeneity between sub-groups | -  -  16.0  8.6 | 1  1  13  2 | -  -  <0.01  <0.01 | 15.3  <0.01  2.8  - |
| **Figure S2**  Day 1  Day 2  Day 3-9  Test for heterogeneity between sub-groups | 6.3  0.0  31.6  3.5 | 8  6  19  2 | <0.01  -  <0.01  0.2 | 0.5  <0.01  0.9  - |
| **Figure S7**  Early  Intermediate  Late  Overall  Test for heterogeneity between sub-groups | 10.8  32.2  31.2  66.2  10.3 | 5  15  15  39  2 | <0.01  <0.01  <0.01  <0.01  <0.01 | 1.3  0.3  0.2  0.3  - |
| **Figure S8**  Cephalosporin  Macrolide  Penicillin  Overall  Test for heterogeneity between sub-groups | 1.0  9.1  10.2  8.9  0.3 | 4  4  17  29  2 | 0.2  <0.01  <0.01  <0.01  0.8 | 0.1  0.3  0.2  0.1  - |
| **Figure S9**  Cephalosporin  Macrolide  Penicillin  Overall  Test for heterogeneity between sub-groups | 0.0  0.0  13.4  16.4  3.2 | 4  4  17  29  2 | -  1.0  <0.01  <0.01  0.2 | <0.01  <0.01  0.4  0.5  - |
| **Figure S10**  Cephalosporin  Lincosamide  Macrolide  Penicillin  Overall  Test for heterogeneity between sub-groups | 0.3  -  142.6  321.6  557.7  7.0 | 3  1  4  11  24  3 | 0.3  0  <0.01  <0.01  <0.01  0.1 | 0.2  <0.01  2.6  2.7  3.0  - |
| **Figure S11**  Cephalosporin  Lincosamide  Macrolide  Penicillin  Overall  Test for heterogeneity between sub-groups | -  -  4.2  17.8  29.3  2.1 | 2  1  4  8  20  3 | -  -  <0.01  <0.01  <0.01  0.5 | 1.0  <0.01  1.3  10.1  3.6  - |
